# Lack of association between HLA and asymptomatic SARS-CoV-2 infection

**DOI:** 10.1101/2023.12.06.23299623

**Authors:** Astrid Marchal, Elizabeth T. Cirulli, Iva Neveux, Evangelos Bellos, Ryan S. Thwaites, Kelly M. Schiabor Barrett, Yu Zhang, Ivana Nemes-Bokun, Mariya Kalinova, Andrew Catchpole, Stuart G. Tangye, András N. Spaan, Justin B. Lack, Jade Ghosn, Charles Burdet, Guy Gorochov, Florence Tubach, Pierre Hausfater, COVID Human Genetic Effort, COVIDeF Study Group, French COVID Cohort Study Group, CoV-Contact Cohort, COVID-STORM Clinicians, COVID Clinicians, Orchestra Working Group, Amsterdam UMC Covid-19 Biobank, NIAID-USUHS COVID Study Group, Clifton L. Dalgard, Shen-Ying Zhang, Qian Zhang, Christopher Chiu, Jacques Fellay, Joseph J. Grzymski, Vanessa Sancho-Shimizu, Laurent Abel, Jean-Laurent Casanova, Aurélie Cobat, Alexandre Bolze

## Abstract

Human genetic studies of critical COVID-19 pneumonia have revealed the essential role of type I interferon-dependent innate immunity to SARS-CoV-2 infection. Conversely, an association between the HLA-B*15:01 allele and asymptomatic SARS-CoV-2 infection in unvaccinated individuals was recently reported, suggesting a contribution of pre-existing T cell-dependent adaptive immunity. We report a lack of association of classical HLA alleles, including HLA-B*15:01, with pre-omicron asymptomatic SARS-CoV-2 infection in unvaccinated participants in a prospective population-based study in the US (191 asymptomatic vs. 945 symptomatic COVID-19 cases). Moreover, we found no such association in the international COVID Human Genetic Effort cohort (206 asymptomatic vs. 574 mild or moderate COVID-19 cases and 1,625 severe or critical COVID-19 cases). Finally, in the Human Challenge Characterisation study, the three HLA-B*15:01 individuals infected with SARS-CoV-2 developed symptoms. As with other acute primary infections, no classical HLA alleles favoring an asymptomatic course of SARS-CoV-2 infection were identified. These findings suggest that memory T-cell immunity to seasonal coronaviruses does not strongly influence the outcome of SARS-CoV-2 infection in unvaccinated individuals.

## Introduction

Primary infection with SARS-CoV-2 underlies a broad spectrum of clinical manifestations in unvaccinated individuals, ranging from silent infection to lethal COVID-19 pneumonia. Rare and common human genetic variants have been associated with hypoxemic COVID-19 pneumonia ^1–5^. Inborn errors of TLR3- and/or TLR7-dependent type I IFN immunity in respiratory epithelial cells and plasmacytoid dendritic cells underlie critical COVID-19 pneumonia in 1-5% of cases ^1,6–8^. Moreover, autoantibodies neutralizing type I interferon (IFN) underlie at least another 15% of cases ^9–11^, further highlighting the key role of type I IFNs in protective immunity to SARS-CoV-2 infection in the respiratory tract. Inborn errors of the OAS-RNase L pathway underlie MIS-C in about 1% of cases ^12^, but the other manifestations of SARS-CoV-2 infection remain unexplained (COVID toes, long COVID etc.). In the large sample study from the COVID-19 Host Genetics Initiative, only one human leukocyte antigen (HLA) class II allele, HLA-DRB1*04:01, has been found to confer a small decrease in the risk of critical COVID-19 (OR = 0.8) ^2^. By contrast, we have documented a stronger association between HLA-A*03:01 and side effects following inoculation with the Pfizer-BioNTech COVID-19 mRNA vaccine ^13^, which was subsequently replicated ^14^.

In this context, in July 2023, an association was reported between the HLA-B*15:01 allele and asymptomatic SARS-CoV-2 infection in unvaccinated individuals ^15^. The OR was 2.40 (95%CI: 1.54–3.64) for heterozygotes, reaching 8.58 (95%CI: 1.74–34.43) in homozygotes. Silent SARS-CoV-2 infection had not hitherto been explicitly studied as a phenotype in large genetic studies. This study further showed that T-cells from HLA-B*15:01 individuals who had not been infected with SARS-CoV-2 recognized a SARS-CoV-2 T-cell epitope by cross-reactivity due to prior exposure to one of two common cold coronaviruses: OC43-CoV or HKU1-CoV^15^. Moreover, more than 100 immunogenic SARS-CoV-2 peptides are highly similar to peptides from at least one human coronavirus (hCoV) presented by a wide range of classical HLA molecules ^16^. We, therefore, tested the hypothesis of an association between HLA alleles and asymptomatic SARS-CoV-2 infection in two large independent cohorts. We aimed: (i) to test the association with HLA-B*15:01 and (ii) to identify additional HLA alleles potentially associated with asymptomatic COVID-19.

## Material and methods

### Cohorts and phenotype information

#### US prospective cohort

Participants in the US prospective cohort came from two studies: the Helix DNA Discovery Project and the Healthy Nevada Project. All the enrolled participants provided written informed consent for participation and were recruited through protocols conforming to local ethics requirements. The Helix DNA Discovery Project was reviewed and approved by the Western Institutional Review Board. For the Healthy Nevada Project (HNP), the University of Nevada, Reno Institutional Review Board approved the study (project 956068-12). The procedures followed were in accordance with ethical standards, and appropriate informed consent was obtained. We performed an online survey that we sent a few times in 2021. The survey takes about 15 minutes to complete and has been published in the past ^13^. We received responses from 8,125 unique Helix DNA Discovery Project participants and 9,315 unique Healthy Nevada Project participants. The participants in this cohort were 18 to 89+ years old, 65% were female, and 85% were of European genetic ancestry. The respondents indicated whether they had been infected and whether they had been vaccinated, as well as information on exposure, reasons for testing, and comorbidities. They rated the severity and duration of their symptoms and disease. They answered questions about 24 specific symptoms known to occur after SARS-CoV-2 infection.

#### CHGE cohort

Since the beginning of the pandemic, the COVID Human Genetic Effort (CHGE) has enrolled more than 10,000 individuals with SARS-CoV-2 infection and broad clinical manifestations from all over the world. All the enrolled participants provided written informed consent for participation and were recruited through protocols conforming to local ethics requirements. For patients enrolled in the French COVID cohort (ClinicalTrials.gov NCT04262921), ethics approval was obtained from the Comité de Protection des Personnes Ile De France VI (ID RCB, 2020-A00256-33) or the Ethics Committee of Erasme Hospital (P2020/203). For participants enrolled in the COV-Contact study (ClinicalTrials.gov NCT04259892), ethics approval was obtained from the CPP IDF VI (ID RCB, 2020-A00280-39). For patients enrolled in the Italian cohort, ethics approval was obtained from the University of Milano-Bicocca School of Medicine, San Gerardo Hospital, Monza–Ethics Committee of the National Institute of Infectious Diseases Lazzaro Spallanzani (84/2020) (Italy), and the Comitato Etico Provinciale (NP 4000–Studio CORONAlab). STORM-Health care workers were enrolled in the STudio OsseRvazionale sullo screening dei lavoratori ospedalieri per COVID-19 (STORM-HCW) study, with approval from the local institutional review board (IRB) obtained on June 18, 2020. Patients and relatives from San Raffaele Hospital (Milan) were enrolled in COVID-BioB/Gene-COVID protocols and, for additional studies, TIGET-06, with the approval of the local ethics committee. Patients and relatives from Rome were enrolled in Protocol no. 50/20 (Tor Vergata University Hospital). Informed consent was obtained from each patient. For the patients enrolled in the COVIDeF Study Group (ClinicalTrials.gov NCT04352348), ethics approval was obtained from the Comité de Protection des Personnes Ile de France XI (ID RCB, 2020-A00754-35). For patients enrolled in Spain, the study was approved by the Committee for Ethical Research of the Infanta Leonor University Hospital, code 008–20; the Committee for Ethical Research of the 12 de Octubre University Hospital, code 16/368; the Bellvitge University Hospital, code PR127/20; the University Hospital of Gran Canaria Dr. Negrín, code 2020–200-1 COVID-19; and the Vall d’Hebron University Hospital, code PR(AMI)388/2016. Anonymized samples were sequenced at the National Institute of Allergy and Infectious Diseases (NIAID) through the Uniformed Services University of the Health Sciences (USUHS)/the American Genome Center (TAGC) under nonhuman subject research conditions; no additional IRB consent was required at the National Institutes of Health (NIH). For patients enrolled in the Swedish COVID cohort, ethics approval was obtained from the Swedish Ethical Review Agency (2020–01911 05).

The physicians classified the patients as follows: i) Critical cases were defined as patients with pneumonia requiring high-flow oxygen (> 6 L/min) and/or requiring admission to the intensive care unit; ii) Severe cases were defined as patients with pneumonia requiring low-flow oxygen (< 6 L/min); iii) Moderate cases were defined as patients with ambulatory pneumonia; iv) Mild cases were defined as pauci-symptomatic patients, with the presence of mild, self-healing symptoms such as cough, fever, body aches, anosmia; and v) Asymptomatic cases were defined as infected individuals with no symptoms. The presence of infection was assessed on the basis of a positive PCR test and/or serological test and/or the presence of typical symptoms such as anosmia or ageusia after exposure to a confirmed COVID-19 case.

#### SARS-CoV-2 Human Challenge Characterisation Study

34 participants seronegative to spike protein were challenged with D614G-containing pre-Alpha SARS-CoV-2, of whom 33 consented for genetic analysis. Additional details on the study design and participants were previously published ^17^. Ethics approval was obtained from the UK Health Research Authority Ad Hoc Specialist Ethics Committee (reference: 20/UK/0002). Written informed consent was obtained from participants before screening and enrollment.

### Sequencing

#### US prospective cohort

DNA samples were sequenced and analyzed at Helix with the Exome+® assay as previously described ^18^. Genotype processing was performed in Hail ^19^.

#### CHGE cohort

Whole-exome (WES) or whole-genome sequencing (WGS) was performed at several sequencing centers, including the Genomics Core Facility of the Imagine Institute (Paris, France), the Yale Center for Genome Analysis (USA), Macrogen (USA), Psomagen (USA), the New-York Genome Center (NY, USA), TAGC (USUHS, Bethesda, USA), MNM Bioscience (Poland), Invitae (San Francisco, USA), the Genomic Sequencing Platform Seqoia (France), the Centre National de Recherche en Génomique Humaine (CNRGH, Evry, France), the Genomics Division-ITER of the Canarian Health System sequencing hub (Canary Islands, Spain), and the AlJalila Genomics Center (Dubai). Libraries for WES were generated with the Twist and Twist Plus Human Core Exome Kit, the xGen Exome Research Panel from Integrated DNA Technologies (IDT; xGen V1 and V2), Agilent SureSelect (Human All Exon V6 and V7) panels, the SeqCap EZ MedExome Kit from Roche, the Nextera Flex for Enrichment-Exome kit, the Illumina TruSeq Exome panel and WES custom target enrichment probes. Massively parallel sequencing was performed on HiSeq 4000, HiSeq 2500, NextSeq 550 or NovaSeq 6000 systems (Illumina).

For principal component analysis (PCA), common variants from the gnomAD v2.1 Exome dataset were jointly genotyped with GATK GenotypeGVCFs. PCA was performed with PLINK v1.9 software on a pruned set of ∼14,600 SNPs not in linkage disequilibrium (maximum r2 value for linkage disequilibrium of 0.4 between pairs of SNPs), with a minor allele frequency (MAF) > 1%, a call rate > 99%, and P value for departure from Hardy–Weinberg equilibrium > 10e−5, as previously described ^20^. The ancestral origin of the patients was further inferred from the PCA, as previously described ^20^.

#### SARS-CoV-2 Human Challenge Characterisation Study

Whole-genome sequencing was performed on Illumina NovaSeq (Novogene Ltd., IK), yielding 150bp paired-end reads. The average depth of coverage was > 50x with a minimum of 31x. PCA and global ancestry inference were performed using Hail according to the protocol described by the gnomAD project ^21^.

### HLA calls / imputation

#### US prospective cohort

HLA types for A, B, C, DPB1, DQA1, DQB1, and DRB1 were imputed with HIBAG using the default recommendations ^22^. Individual genotypes were imputed with the model for the most appropriate genetic ancestry for each individual. Probabilities greater than 0.5 were used for genotype calling.

#### CHGE cohort

Classical class I and class II HLA alleles were typed from the raw WES or WGS reads with HLA*LA software [10], which uses a linear projection method to align reads to a population reference graph and enables high HLA typing accuracy from WES or WGS data.

#### SARS-CoV-2 Human Challenge Characterisation Study

HLA alleles were typed from raw WGS reads with HLA*LA software at G group resolution. Only HLA calls with a posterior probability of 100% and a minimum coverage of 20x were retained in the analysis. At the B locus, all individual calls fulfilled these filtering criteria at 2-field resolution.

These tools have been validated for their accuracy to call HLA alleles at 2-field resolution, particularly in populations of European ancestry. For example, the HIBAG HLA calls made at Helix for 7 genes in 7 European ancestry Coriell samples showed 99% concordance with the known HLA calls for these individuals. Differences caused by HLA allele calling should mostly be limited to rare HLA types and populations with poor imputation references.

### HLA-WAS

We used Regenie ^23^ for the genetic analysis. In brief, this method builds a whole-genome regression model based on common variants to account for the effects of relatedness and population stratification; it also accounts for situations in which there is an extreme case-control imbalance likely to lead to test statistic inflation with other analysis methods. We used the approximate Firth p value when the logistic regression p value was below 0.01. The covariates included were age group, sex and the first five principal components.

For the US prospective cohort, a representative set of 184,445 coding and noncoding LD-pruned, high-quality common variants were identified for the construction of PCs and the whole-genome regression model, as previously described ^18^. PCs were calculated within the European group. For CHGE, the set of ∼14,600 SNPs used for PCA within the European group was used for the whole-genome regression model.

### Meta-analysis

Results were combined by inverse variance-weighted fixed-effects meta-analysis with METAL ^24^. Effect was provided as the BETA value and the STDERR was provided as the SE.

### Power calculation

We estimated the power required to detect an effect similar to that reported by Augusto, Murdolo & Chatzileontiadou et al. with the Genetic Association Study (GAS) Power Calculator, which uses a method derived from the CaTS power calculator for two-stage association studies^25^. The parameters used were: HLA-B*15:01 frequency: 0.05; prevalence of asymptomatic infection: 0.1; Dominant inheritance model; p-value threshold = 0.05; numbers of cases and controls according to the third definition in both cohorts.

### Serology

Plasma IgG titres were determined using MesoScale Discovery Coronavirus panel 2 plates on a SQ120 instrument. Binding titres given as arbitrary units per milliliter (AU/ml) based on a kit-provided human plasma standard curve.

## Results

### US prospective cohort description

We first conducted an HLA-wide association study (HLA-WAS) in a prospective population-based US cohort (**Figure 1**). Participants were either part of the Helix DNA Discovery Project or the Healthy Nevada project, and were recruited before the start of the COVID-19 pandemic. All participants underwent Exome+^®^ sequencing, which targets the exome and a few hundred thousand non-exonic common SNPs, providing a backbone for imputation of the most common SNPs in the genome. HLA alleles were called for seven genes (HLA -A, -B, -C, -DPB1, -DQA1, -DQB1, and -DRB1) with HIBAG ^22^. The 17,434 adults who responded to at least one of the COVID-19 infection and vaccination surveys sent in 2021 included 1,680 participants reporting SARS-CoV-2 infection while unvaccinated. A continuous spectrum of symptoms, duration of illness was reported following SARS-CoV-2 infection (**Figure S1**). The most common symptoms were muscle and body aches, and a cough (**Figure S1A**). No symptoms at all were reported by 5.1% of individuals (n=86), whereas 5.3% of the infected participants required hospitalization with or without oxygen therapy (n=58) or were admitted to the intensive care unit (n=31) (**Figure S1B**).

**Figure 1:**
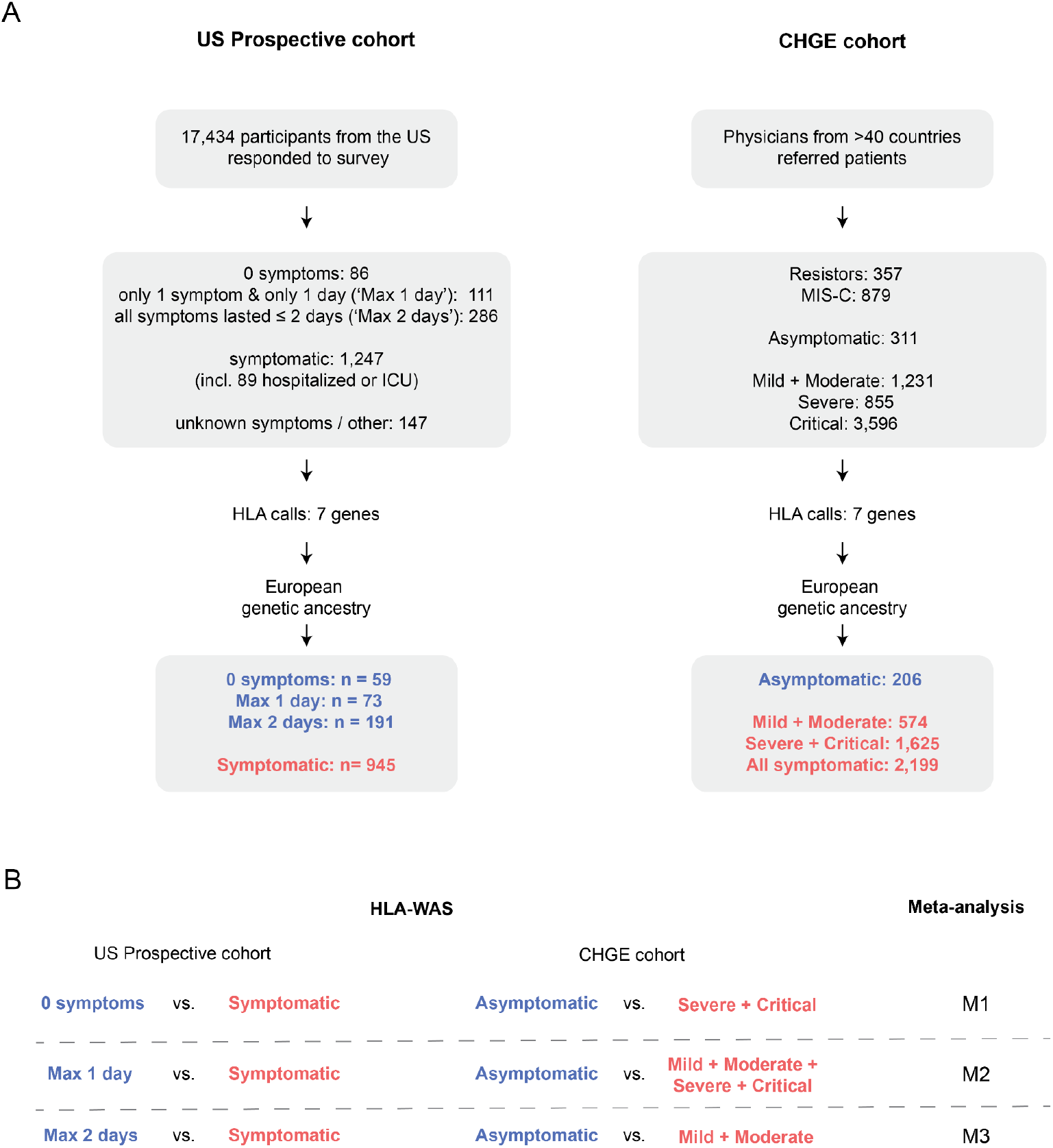
Study design. A: Description of the two cohorts and definitions of asymptomatic and symptomatic cases. B: List of HLA-wide association studies and meta-analyses performed.

### HLA-wide association in the US prospective cohort

We tested the hypothesis that HLA alleles play an important role in the early response to SARS-CoV-2 by considering three case definitions for the asymptomatic cases (**Figure 1A**): 1) ‘0 symptoms’ was a stringent definition of asymptomatic as a total absence of symptoms (n=86); 2) ‘Max 1 day’ was a definition of asymptomatic in which the presence of one symptom for no more than one day was tolerated (n=111). This definition was used to increase the power for detection of an association by enlarging the ‘asymptomatic’ group whilst still identifying individuals who cleared the virus quickly and efficiently; 3) ‘Max 2 days’ was a definition as close as possible to that used by Augusto, Murdolo & Chatzileontiadou et al., considering participants to be asymptomatic if none of their symptoms lasted three days or more, and if the reason for testing was unrelated to symptoms (n=286) ^15^. We used only one definition for controls (individuals with symptoms lasting at least three days). The control group included all individuals admitted to the ICU or the hospital and anyone reporting symptoms of at least three days’ duration with some impact on their daily routine (n=1,247). For the HLA-WAS, we restricted the analysis to individuals with a genetically inferred European ancestry (**Figure S1C-D**), leading to a total of 59 to 191 asymptomatic cases and of 945 symptomatic controls (**Figure 1A**). Age and sex distribution are shown in **Table S1**. The association test was performed with Regenie ^23^ under a dominant inheritance model, with age, sex and the first five principal components as covariates (see **Supplemental methods**). The risk of detecting false positive associations was decreased by limiting the analysis to the 105 HLA alleles with an allele frequency of at least 1% in this cohort. No statistically significant associations (at a corrected threshold of p<0.00047) were found with any of the three phenotype definitions (**Table 1**, **Tables S2-4**). The top-ranked HLA allele was DRB1*16:01, which was depleted in asymptomatic individuals, with the strongest effect being observed in the ‘Max 2 days’ group of asymptomatic patients (OR [95%CI] = 0.06 [0 – 1.5], p=0.004, **Table 1**).

**Table 1:** Top-ranked alleles in the HLA-WAS on the US prospective cohort.

| Allele | OR [95% CI]<sup>a</sup> | p value | AF<sup>b</sup> | Asymptomatic definition<sup>c</sup> |
| --- | --- | --- | --- | --- |
| DRB1*16:01 | 0.06 [0.002-1.5] | 0.004 | 0.012 | Max 2 days |
| A*32:01 | 2.19 [1.3-3.8] | 0.008 | 0.035 | Max 2 days |
| B*07:02 | 1.58 [1.1-2.3] | 0.016 | 0.13 | Max 2 days |
| C*07:02 | 1.53 [1.1-2.2] | 0.020 | 0.14 | Max 2 days |
| DQB1*06:02 | 1.52 [1.1-2.2] | 0.025 | 0.14 | Max 2 days |
| DQB1*05:02 | 0.29 [0.1-1.1] | 0.037 | 0.015 | Max 2 days |

<sup>a</sup> Odds ratio (OR) of being asymptomatic, i.e. $OR > 1$ indicates that the allele is more frequent in asymptomatic individuals. CI: confidence interval.
<sup>b</sup> Allele frequency (AF) is based on the frequency of the allele in the US prospective cohort ‘Max 2 days’ analysis because this analysis included the largest numbers of cases and controls.
<sup>c</sup> For each allele, the top-ranked result across the three asymptomatic definitions in the US prospective cohort is given.

### HLA-wide association in the CHGE cohort

We next studied patients recruited by the physicians of the international CHGE consortium. These physicians classified participants with SARS-CoV-2 infections according to acute disease severity: asymptomatic, mild, moderate, severe, or critical (**Figure S2A**). Whole-exome or whole-genome sequencing data were available for 7,229 participants and HLA alleles were typed with HLA*LA ^26^. In this HLA-WAS, we compared the patients classified as ‘asymptomatic’ by the clinicians (n=311) with those in three sets of symptomatic controls: 1) the patients with the most extreme symptoms requiring hospitalization and oxygen supplementation (i.e. those with a severe or critical form of the disease, n=4,451); 2) all symptomatic patients, whatever their acute disease severity (i.e. mild, moderate, severe or critical, n=5,682); and 3) symptomatic patients not requiring oxygen supplementation (i.e. mild and moderate patients only, n=1,231); this last group of symptomatic patients is the most similar to the symptomatic patients groups of the US prospective cohort and the study by Augusto, Murdolo & Chatzileontiadou et al. (**Figure 1**). We restricted the analysis to individuals of European genetic ancestry and the final study population comprised 206 asymptomatic cases, 1,625 patients with severe or critical disease and 574 patients with mild or moderate disease **(Figure S2B-C**). Age and sex distribution are shown in **Table S1**. Analyses were also performed under a dominant inheritance model with age group, sex and the first five principal components as covariates. This analysis was performed with Regenie and was limited to the 117 HLA alleles with an allele frequency of at least 1% in this cohort. No statistically significant association (at a corrected threshold of p<0.00043) was identified in the HLA-WAS, regardless of the definition of symptomatic patients used (**Tables S5-S7**). The top-ranked HLA allele found to be enriched in asymptomatic individuals was HLA-B*40:02, for which the strongest effect was observed in comparison with the group of symptomatic patients with severe or critical disease (OR [95%CI] = 3.4 [1.5 – 7.7], p=0.005, **Table 2**).

**Table 2:** Top-ranked alleles in the HLA-WAS on the CHGE European cohort.

| Allele | OR [95% CI]<sup>a</sup> | p value | AF<sup>b</sup> | Controls used<sup>c</sup> |
| --- | --- | --- | --- | --- |
| B*40:02 | 3.42 [1.5-7.7] | 0.005 | 0.016 | Severe + Critical |
| DPB1*01:01 | 0.28 [0.1-0.8] | 0.007 | 0.042 | Severe + Critical |
| A*23:01 | 2.5 [1.2-5.0] | 0.010 | 0.023 | Mild + Moderate |
| B*49:01 | 2.26 [1.2-4.3] | 0.014 | 0.031 | Mild + Moderate |
| A*03:01 | 1.66 [1.1-2.5] | 0.019 | 0.12 | Severe + Critical |
| DQA1*01:02 | 1.54 [1.1-2.2] | 0.022 | 0.18 | Mild + Moderate |
| A*30:02 | 2.46 [1.1-5.6] | 0.031 | 0.019 | Mild + Moderate |
| B*57:01 | 0.47 [0.2-1.0] | 0.047 | 0.027 | Mild + Moderate |
| A*68:02 | 3.53 [1.0-12.2] | 0.047 | 0.01 | Mild + Moderate |
| DPB1*03:01 | 0.65 [0.4-1.0] | 0.049 | 0.093 | Mild + Moderate |

<sup>a</sup> Odds ratio (OR) of being asymptomatic, i.e. OR>1 indicates that the allele is more frequent in asymptomatic individuals. CI: confidence interval.
<sup>b</sup> Allele frequency (AF) is based on the frequency of the allele in the CHGE European cohort 'All' analysis, which included the largest numbers of cases and controls.
<sup>c</sup> For each allele, the top-ranked result across three sets of symptomatic patients in the CHGE European sample is given.

### HLA-wide meta-analysis

We then performed three meta-analyses, denoted M1, M2, M3 (**Figure 1B**), combining the results from our two independent cohorts with METAL ^24^. The first used the strictest definitions for the groups: the HLA-WAS with the ‘0 symptoms’ group of asymptomatic patients in the US prospective cohort and the HLA-WAS limited to patients with severe and critical disease only in the CHGE cohort (**Table S8**). The second meta-analysis combined the HLA-WAS with the ‘Max 1 day’ definition of asymptomatic patients for the US prospective cohort (0 symptoms or 1 symptom for 1 day) with the HLA-WAS with all symptomatic cases from the CHGE (**Table S9**). The final meta-analysis used the results for the asymptomatic and symptomatic groups most closely resembling those of the study by Augusto, Murdolo & Chatzileontiadou et al. (**Table S10**). The meta-analyses detected no statistically significant associations (at a corrected threshold of p<0.00047) between HLA alleles and asymptomatic SARS-CoV-2 infection (**Table 3**, **Tables S8-10**). The top-ranked HLA allele was HLA-B*40:02 (p-value = 0.0008), for which enrichment was observed in asymptomatic individuals relative to symptomatic individuals in both cohorts and in the meta-analysis based on the strictest definitions.

**Table 3:** Top-ranked alleles in the meta-analyses and corresponding results in the US prospective and CHGE cohorts.

| Allele | Meta-analysis |  |  | US Prospective Cohort<sup>b</sup> |  | CHGE Cohort<sup>b</sup> |  |
| --- | --- | --- | --- | --- | --- | --- | --- |
|  | Meta-analysis number | OR [95% CI]<sup>a</sup> | p value | OR [95% CI]<sup>a</sup> | p value | OR [95% CI]<sup>a</sup> | p value |
| B*40:02 | M1 | 3.51 [1.7-7.3] | 0.0008 | 4.05 [0.7-24.6] | 0.128 | 3.42 [1.5-7.7] | 0.005 |
| DPB1*01:01 | M2 | 0.43 [0.3-0.7] | 0.0015 | 0.43 [0.2-1.0] | 0.058 | 0.43 [0.2-0.8] | 0.010 |
| DQA1*01:02 | M3 | 1.41 [1.1-1.8] | 0.007 | 1.31 [0.9-1.8] | 0.119 | 1.54 [1.1-2.2] | 0.022 |
| A*23:01 | M2 | 2.22 [1.2-4.0] | 0.007 | 2.14 [0.7-6.6] | 0.186 | 2.25 [1.1-4.4] | 0.019 |
| DQB1*06:02 | M3 | 1.46 [1.1-2.0] | 0.013 | 1.52 [1.1-2.2] | 0.025 | 1.33 [0.8-2.2] | 0.276 |
| C*03:03 | M1 | 0.52 [0.3-0.9] | 0.015 | 0.48 [0.2-1.2] | 0.132 | 0.54 [0.3-1.0] | 0.055 |
| B*49:01 | M3 | 1.96 [1.1-3.4] | 0.019 | 1.29 [0.4-3.9] | 0.657 | 2.26 [1.2-4.3] | 0.014 |
| B*07:02 | M3 | 1.42 [1.1-1.9] | 0.021 | 1.58 [1.1-2.3] | 0.016 | 1.16 [0.7-1.9] | 0.554 |
| DRB1*15:01 | M3 | 1.38 [1.0-1.9] | 0.037 | 1.42 [1-2.1] | 0.075 | 1.32 [0.8-2.2] | 0.264 |

<sup>a</sup> Odds ratio (OR) of being asymptomatic, i.e. OR>1 indicates that the allele is more frequent in asymptomatic individuals. CI: confidence interval.
<sup>b</sup> For each allele, ORs and p values obtained in the US prospective and CHGE cohorts with asymptomatic or controls definitions used in the corresponding meta-analysis are given.

### Lack of replication for HLA-B*15:01

An analysis focusing on HLA-B*15:01 did not replicate the association between HLA-B*15:01 and asymptomatic SARS-CoV-2 infection (**Figure 2A-C**) despite being well powered (>95%) to detect an effect similar to that reported by Augusto, Murdolo & Chatzileontiadou et al. (OR of 2.40 for enrichment in asymptomatic vs. symptomatic patients, p=5.67x10^-5^) (**Figure S3**). We further estimated the frequency of HLA-B*15:01 in various groups of patients of the CHGE consortium, including children with SARS-CoV-2 infection complicated by multisystem inflammatory syndrome (classified as MIS-C) and individuals with high levels of exposure who never tested positive (classified as ‘resistors’) ^12,27^. This frequency ranged from 2.4% in asymptomatic individuals to 6.0% in resistors (**Figure 2C-D**). We also looked at individuals from non-European genetic ancestries. Similarly, we found no difference in frequency between asymptomatic and symptomatic individuals (**Figure 2E** **and Table S11**). Overall, no enrichment in the HLA-B*15:01 allele was observed among asymptomatic individuals in our US population-based prospective cohort, or in the international CHGE cohort.

**Figure 2:**
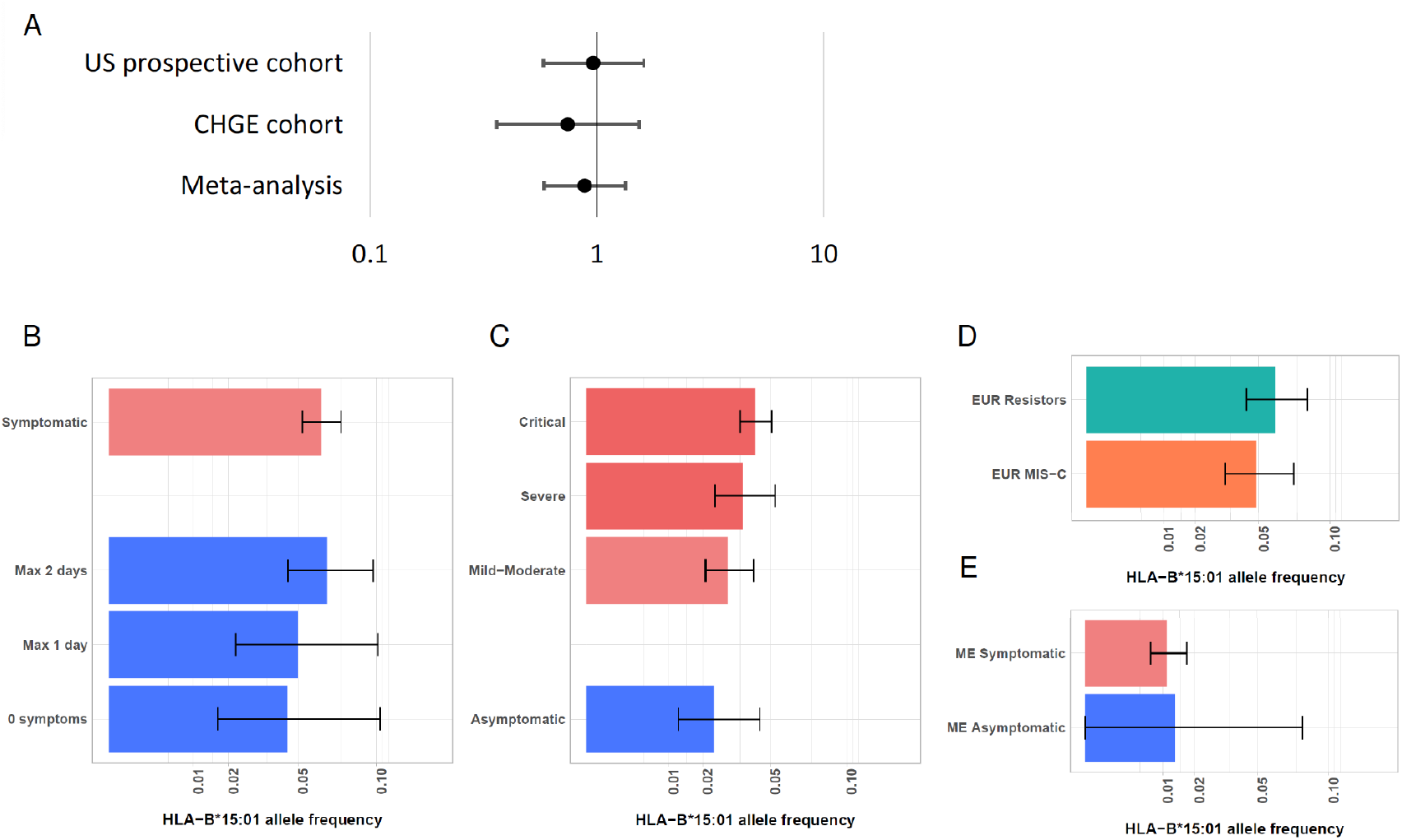
HLA-B*15:01 is not enriched in asymptomatic cases. A: Odds Ratio (OR) and 95% confidence intervals (CI) for the association of HLA-B*15:01 with asymptomatic SARS-CoV-2 infection in both cohorts and in the meta-analysis. B: Allele frequency and 95% confidence intervals in the US prospective cohort subgroups. C: Allele frequency and 95% CI in the CHGE European sample. D: Allele frequency and 95% CI in individuals highly exposed to SARS-CoV-2 who never tested positive (‘Resistors’, n=291) and children with SARS-CoV-2 infection complicated by multisystem inflammatory syndrome (‘MIS-C’, n=235) from the European CHGE sample. E: Allele frequency and 95% CI in Middle Eastern (ME) individuals from the CHGE cohort (Symptomatic, n=895; Asymptomatic, n=37).

### Symptoms and serology for participants with HLA-B*15:01 in the SARS-CoV-2 Human Challenge Characterisation Study

The mechanism proposed as an explanation for the association between HLA-B*15:01 and asymptomatic SARS-CoV-2 infection was pre-existing immunity, probably due to prior infection with OC43-CoV or HKU1-CoV ^15^. Unfortunately, no serological data were available for the HLA-B*15:01 carriers in the US prospective and the CHGE cohorts. We tested the hypothesis that the lack of association in our study was due to an absence of prior infection with OC43-CoV or HKU1-CoV by examining the data for the SARS-CoV-2 Human Challenge Characterisation Study (ClinicalTrials.gov identifier NCT04865237; funder, UK Vaccine Taskforce), in which 34 participants seronegative to spike protein were challenged with D614G-containing pre-Alpha SARS-CoV-2, of whom 33 consented for genetic analysis ^17^. Serological data, history of prior infections with other coronaviruses and genetic data were available, together with infection status and data concerning the recorded symptoms. HLA alleles were called with HLA*LA from whole-genome sequences obtained from the participants. Three of the 17 infected participants (positive test result) carried an HLA-B*15:01 allele, as well as three of the 16 who stayed uninfected. Only one of the 17 infected participants was fully asymptomatic and this participant did not carry the HLA-B*15:01 allele. The three infected participants with an HLA-B*15:01 allele were symptomatic (**Figure 3**), despite evidence of prior exposure to OC43-CoV and HKU1-CoV (**Figure S4**). Thus, prior exposure to a coronavirus did not prevent the HLA-B*15:01 carriers from developing symptoms following SARS-CoV-2 infection.

**Figure 3:**
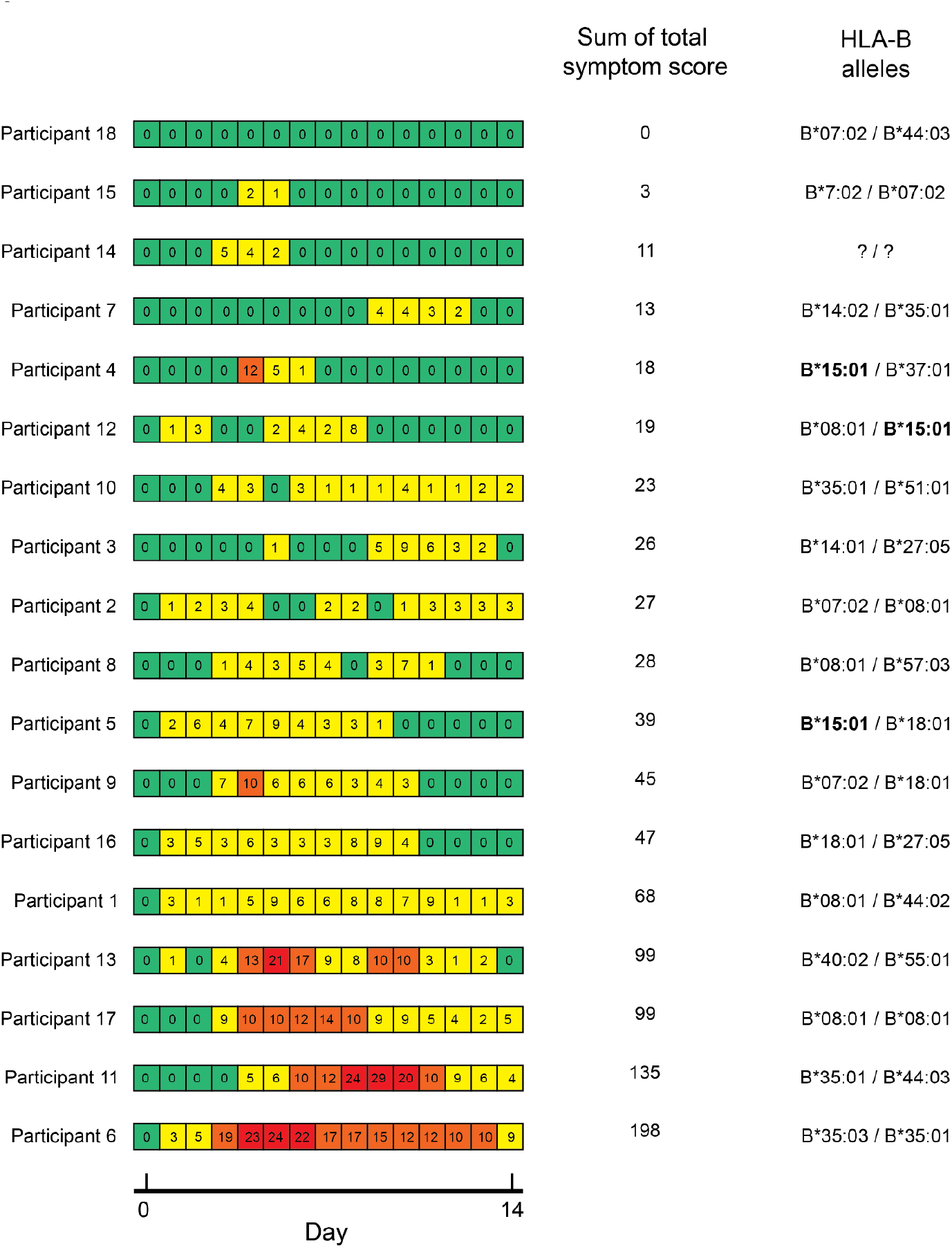
HLA-B*15:01 in the SARS-CoV-2 Human Challenge Characterisation Study. Symptoms and HLA-B genotypes for 18 infected participants. Daily total symptom score was calculated using self-reported symptom diaries three times daily. Daily total symptom scores are displayed in the heatmap, ranging from green (no symptoms) to red (highest symptom score). The heatmap is derived from figure 2 in Zhou J. et al, *Lancet Microbe* (2023).

## Discussion

Our analyses identified no associations between classical HLA alleles and asymptomatic SARS-CoV-2 infection. In particular, we did not replicate the previously reported association between HLA-B*15:01 and asymptomatic SARS-CoV-2 infection. Another recent study in a Spanish cohort found no associations between classical HLA alleles and asymptomatic SARS-CoV-2 infection ^28^. One possible explanation for the difference in results regarding HLA-B*15:01 is that the studies analyzed different groups of individuals living in different environments. However, the US prospective cohort we analyzed has many features in common with the cohort analyzed by Augusto, Murdolo & Chatzileontiadou et al.: specifically, the participants were from the US, with a slight bias towards women, the phenotype was assessed on the basis of self-reported surveys at multiple time points during the pandemic before summer 2021 (before the SARS-CoV-2 Delta variant became dominant in the US ^29^). The percentage of individuals self-reporting asymptomatic infection were similar between the two, as were the rates of each symptom. Alternatively, the difference in results may reflect differences in the handling of potential population stratification. Augusto, Murdolo & Chatzileontiadou et al. did not consider population structure in their study on bone marrow donors, probably because no genetic information outside of the HLA region was available, whereas we accounted for population structure by restricting our analysis to those of European ancestry and including the first five principal components as covariates in our regression model. The highly polymorphic nature of the HLA region and the differences in allele frequencies between human sub-populations contribute to a high risk of false-positive results in association analyses. The frequency of HLA-B*15:01 is known to vary across continents, between continental populations within the US (**Figure S5A**) and even between European countries (**Figure S5B**). Population stratification may, thus, have played a confounding role in the study by Augusto, Murdolo & Chatzileontiadou et al.

Overall, the absence of an association between classical HLA alleles and asymptomatic SARS-CoV-2 infection is consistent with the modest impact of HLA variation on severe or critical COVID-19 ^30^. Most genetic and immunological studies of severe or critical COVID-19 pneumonia in unvaccinated individuals have implicated type I IFNs, suggesting that intrinsic and innate immunity play a more crucial role than adaptive immunity in the early response to SARS-CoV-2. Pre-existing immunity due to prior infections with common cold coronaviruses may help to prevent the development of symptoms following SARS-CoV-2 infection, but our results suggest that either (i) pre-existing CD4 and CD8 T-cell immunity may not play an important role, or (ii) pre-existing immunity is not stronger for individuals with a particular HLA allele than for those with any other HLA allele. This result is also consistent with the absence of any strong association between HLA alleles and clinical outcomes during the acute phase for the other primary viral infections studied to date ^31–33^. By contrast, HLA alleles have been associated with multiple clinical or laboratory outcomes during chronic infections, including viral (e.g., HIV, HBV, HCV), mycobacterial (e.g., leprosy) and protozoan infections ^33–36^. HLA alleles are also known to be associated with adaptive immune responses to vaccinations ^13,37,38^. Our findings suggest that memory T-cell immunity to seasonal coronaviruses does not strongly influence the outcome of SARS-CoV-2 infection in unvaccinated individuals.

## Declaration of interests

E.T.C., K.M.S.B., and A.B. are employees of Helix.

## Acknowledgements

Funding was provided to the Desert Research Institute (DRI) by the Nevada Governor’s Office of Economic Development. Funding was provided by Renown Health and the Renown Health Foundation.

The Laboratory of Human Genetics of Infectious Diseases is supported by the Howard Hughes Medical Institute, the Rockefeller University, the St. Giles Foundation, the National Institutes of Health (NIH) (R01AI63029), the National Center for Advancing Translational Sciences (NCATS), NIH Clinical and Translational Science Award (CTSA) program (UL1 TR001866), the Yale Center for Mendelian Genomics and the GSP Coordinating Center funded by the National Human Genome Research Institute (NHGRI) (UM1HG006504 and U24HG008956), the Yale High Performance Computing Center (S10OD018521), the Fisher Center for Alzheimer’s Research Foundation, the JPB Foundation, the Meyer Foundation, the French National Research Agency (ANR) under the “Investments for the Future” program (ANR-10-IAHU-01), the Integrative Biology of Emerging Infectious Diseases Laboratory of Excellence (ANR-10-LABX-62-IBEID), the French Foundation for Medical Research (FRM) (EQU201903007798), the ANRS-COV05, ANR GENVIR (ANR-20-CE93-003), and ANR AI2D (ANR-22-CE15-0046) projects, the ANR-RHU program (ANR-21-RHUS-08), the European Union’s Horizon 2020 research and innovation program under grant agreement No. 824110 (EASI-genomics), the HORIZON-HLTH-2021-DISEASE-04 program under grant agreement 01057100 (UNDINE), the Square Foundation, Grandir – Fonds de solidarité pour l’enfance, Fondation du Souffle, the SCOR Corporate Foundation for Science, *William E. Ford, General Atlantic’s Chairman and Chief Executive Officer, Gabriel Caillaux, General Atlantic’s Co-President, Managing Director and Head of business in EMEA, and the General Atlantic Foundation*, the Battersea & Bowery Advisory Group, The French Ministry of Higher Education, Research, and Innovation (MESRI-COVID-19), Institut National de la Santé et de la Recherche Médicale (INSERM), REACTing-INSERM and the University of Paris Cité. A.N.S. is supported by European Union’s Horizon Health research and innovation program under grant agreement No 101057100, project UNDINE.

The study was supported by the ORCHESTRA project, which has received funding from the European Union’s Horizon 2020 research and innovation program under grant agreement No 10101616. The French COVID Cohort study group was sponsored by INSERM and supported by the REACTing consortium and by a grant from the French Ministry of Health (Grant PHRC 20-0424). The Cov-Contact Cohort was supported by the REACTing consortium, the French Ministry of Health, and the European Commission (Grant RECOVER WP 6). The COVIDeF study group was supported by the French Ministry of Health, Fondation AP-HP et Programme Hospitalier de Recherche Clinique (PHRC COVID-19-20-0048). Y.Z. and H.C.S. are supported by the Intramural Research Program of the National Institute of Allergy and Infectious Diseases, NIH. G.N. and A.N. are supported by Regione Lazio (Research Group Projects 2020) No. A0375-2020-36663, GecoBiomark. This project has received funding from the European Research Council (ERC) under the European Union’s Horizon 2020 research and innovation program (grant agreement no. 948959). This work is supported by the Swiss National Science Foundation (grant # 310030L_197721 to JF). This work is supported by ERN-RITA. The Canarian Sequencing Hub is funded by Instituto de Salud Carlos III (COV20_01333, and COV20_01334, and PI20/00876) and Spanish Ministry of Science and Innovation (RTC-2017-6471-1; AEI/FEDER, UE), co-financed by the European Regional Development Funds, “A way of making Europe” from the European Union, and Cabildo Insular de Tenerife (CGIEU0000219140 and “Apuestas científicas del ITER para colaborar en la lucha contra la COVID-19”). This work was funded, at least in part, by grants AJF202019 and AJF20259 from Al Jalila Foundation, Dubai, United Arab Emirates. Sample processing at IrsiCaixa was possible thanks to the crowdfunding initiative YoMeCorono.

We thank I Erkizia, E Grau, M Massanella, and J Guitart from the IrsiCaixa and Hospital Germans Trias i Pujol (Badalona, Spain) for sample collection, handling and processing.

We thank the patients and their families for agreeing to participate in our research.

We thank Helen C. Su from the NIAID (Bethesda, USA) and all members of the consortia listed below:

***Members of COVID Human Genetic Effort*:** Laurent Abel^1^, Alessandro Aiuti^2^, Saleh Al-Muhsen^3^, Fahd Al-Mulla^4^, Ali Amara^5^, Mark S. Anderson^6^, Evangelos Andreakos^7^, Andrés A. Arias^8^, Lisa M. Arkin^9^, Hagit Baris Feldman^10^, Paul Bastard^1^, Alexandre Belot^11^, Catherine M. Biggs^12^, Dusan Bogunovic^13^, Alexandre Bolze^14^, Anastasiia Bondarenko^15^, Alessandro Borghesi^16^, Ahmed A. Bousfiha^17^, Petter Brodin^18^, Yenan Bryceson^19^, Manish J. Butte^20^, Jean-Laurent Casanova^21^, Giorgio Casari^22^, John Christodoulou^23^, Aurélie Cobat^1^, Roger Colobran^24^, Antonio Condino-Neto^25^, Stefan N. Constantinescu^26^, Megan A. Cooper^27^, Clifton L. Dalgard^28^, Murkesh Desai^29^, Beth A. Drolet^30^, Xavier Duval^31^, Jamila El Baghdadi^32^, Philippine Eloy^33^, Sara Espinosa-Padilla^34^, Jacques Fellay^35^, Carlos Flores^36^, José Luis Franco^37^, Antoine Froidure^38^, Guy Gorochov^39^, Peter K. Gregersen^40^, Bodo Grimbacher^41^, Filomeen Haerynck^42^, David Hagin^43^, Rabih Halwani^44^, Lennart Hammarström^45^, James R. Heath^46^, Elena W.Y. Hsieh^47^, Eystein Husebye^48^, Kohsuke Imai^49^, Yuval Itan^50^, Emmanuelle Jouanguy^1^, Elżbieta Kaja^51^, Timokratis Karamitros^52^, Kai Kisand^53^, Cheng-Lung Ku^54^, Yu-Lung Lau^55^, Yun Ling^56^, Carrie L. Lucas^57^, Tom Maniatis^58^, Davood Mansouri^59^, László Maródi^60^, France Mentré^32^, Isabelle Meyts^61^, Joshua D. Milner^62^, Kristina Mironska^63^, Trine H. Mogensen^64^, Tomohiro Morio^65^, Lisa F.P. Ng^66^, Luigi D. Notarangelo^67^, Antonio Novelli^68^, Giuseppe Novelli^69^, Cliona O’Farrelly^70^, Satoshi Okada^71^, Keisuke Okamoto^72^, Tayfun Ozcelik^73^, Qiang Pan-Hammarström^45^, Jean W. Pape^74^, Rebeca Perez de Diego^75^, Jordi Perez-Tur^76^, David S. Perlin^77^, Graziano Pesole^78^, Anna M. Planas^79^, Carolina Prando^80^, Aurora Pujol^81^, Anne Puel^1^, Lluis Quintana-Murci^82^, Sathishkumar Ramaswamy^83^, Laurent Renia^66^, Igor Resnick^84^, Carlos Rodríguez-Gallego^85^, Vanessa Sancho-Shimizu^86^, Anna Sediva^87^, Mikko R.J. Seppänen^88^, Mohammad Shahrooei^89^, Anna Shcherbina^90^, Ondrej Slaby^91^, Andrew L. Snow^92^, Pere Soler-Palacín^93^, Vassili Soumelis^94^, András N. Spaan^95^, Ivan Tancevski^96^, Stuart G. Tangye^97^, Ahmad Abou Tayoun^83^, Şehime Gülsün Temel^98^, Christian Thorball^99^, Pierre Tiberghien^100^, Sophie Trouillet-Assant^101^, Stuart E. Turvey^102^, K M Furkan Uddin^103^, Mohammed J. Uddin^104^, Diederik van de Beek^105^, Donald C. Vinh^106^, Horst von Bernuth^107^, Joost Wauters^108^, Mayana Zatz^109^, Pawel Zawadzki^110^, Qian Zhang^1^, Shen-Ying Zhang^1^

^1^Laboratory of Human Genetics of Infectious Diseases, Necker Branch, INSERM U1163, Necker Hospital for Sick Children, Paris, France; Paris Cité University, Imagine Institute, Paris, France; St Giles Laboratory of Human Genetics of Infectious Diseases, Rockefeller Branch, Rockefeller University, New York, NY, USA, ^2^San Raffaele Telethon Institute for Gene Therapy, IRCCS Ospedale San Raffaele, and Vita Salute San Raffaele University, Milan, Italy, ^3^Immunology Research Lab, Department of Pediatrics, College of Medicine, King Saud University, Riyadh, Saudi Arabia, ^4^Dasman Diabetes Institute, Department of Genetics and Bioinformatics, Dasman, Kuwait, ^5^Laboratory of Genomes & Cell Biology of Disease, INSERM U944, CNRS UMR 7212, Université de Paris, Institut de Recherche Saint-Louis, Hôpital Saint-Louis, Paris, France, ^6^Diabetes Center, University of California San Francisco, San Francisco, CA, USA, ^7^Laboratory of Immunobiology, Center for Clinical, Experimental Surgery and Translational Research, Biomedical Research Foundation of the Academy of Athens, Athens, Greece, ^8^St Giles Laboratory of Human Genetics of Infectious Diseases, Rockefeller Branch, The Rockefeller University, New York, NY, USA; Primary Immunodeficiencies Group, Department of Microbiology and Parasitology, School of Medicine, University of Antioquia, Medellín, Colombia; School of Microbiology, University of Antioquia UdeA, Medellín, Colombia, ^9^Department of Dermatology, School of Medicine and Public Health, University of Wisconsin-Madison, Madison, WI, USA, ^10^The Genetics Institute, Tel Aviv Sourasky Medical Center and Sackler Faculty of Medicine, Tel Aviv University, Tel Aviv, Israel, ^11^Pediatric Nephrology, Rheumatology, Dermatology, HFME, Hospices Civils de Lyon, National Referee Centre RAISE, and INSERM U1111, Université de Lyon, Lyon, France, ^12^Department of Pediatrics, BC Children’s and St Paul’s Hospitals, University of British Columbia, Vancouver, BC, Canada, ^13^Icahn School of Medicine at Mount Sinai, New York, NY, USA, ^14^Helix, San Mateo, CA, USA, ^15^International European University, Kyiv, Ukraine, ^16^Neonatal Intensive Care Unit, Fondazione IRCCS Policlinico San Matteo, Pavia, Italy, ^17^Clinical Immunology Unit, Department of Pediatric Infectious Disease, CHU Ibn Rushd and LICIA, Laboratoire d’Immunologie Clinique, Inflammation et Allergie, Faculty of Medicine and Pharmacy, Hassan II University, Casablanca, Morocco, ^18^SciLifeLab, Department Of Women’s and Children’s Health, Karolinska Institutet, Stockholm, Sweden, ^19^Department of Medicine, Center for Hematology and Regenerative Medicine, Karolinska Institutet, Stockholm, Sweden, ^20^Division of Immunology, Allergy, and Rheumatology, Department of Pediatrics and the Department of Microbiology, Immunology, and Molecular Genetics, University of California, Los Angeles, CA, USA, ^21^The Rockefeller University & Howard Hughes Medical Institute, New York, NY, USA; Necker Hospital for Sick Children & INSERM, Paris, France, ^22^Clinical Genomics, IRCCS San Raffaele Scientific Institute and Vita-Salute San Raffaele University, Milan, Italy, ^23^Murdoch Children’s Research Institute and Department of Paediatrics, University of Melbourne, Melbourne, VIC, Australia, ^24^Immunology Division, Genetics Department, Hospital Universitari Vall d’Hebron, Vall d’Hebron Research Institute, Vall d’Hebron Barcelona Hospital Campus, UAB, Barcelona, Catalonia, Spain, ^25^Department of Immunology, Institute of Biomedical Sciences, University of São Paulo, São Paulo, Brazil, ^26^de Duve Institute and Ludwig Cancer Research, Brussels, Belgium, ^27^Washington University School of Medicine, St Louis, MO, USA, ^28^Department of Anatomy, Physiology & Genetics, Uniformed Services University of the Health Sciences, Bethesda, MD, USA, ^29^Bai Jerbai Wadia Hospital for Children, Mumbai, India, ^30^School of Medicine and Public Health, University of Wisconsin, Madison, WI, USA, ^31^Université de Paris, IAME UMR-S 1137, INSERM, Paris, France; Inserm CIC 1425, Paris, France, ^32^Genetics Unit, Military Hospital Mohamed V, Rabat, Morocco, ^33^Hôpital Bichat, Paris, France, ^34^Instituto Nacional de Pediatria (National Institute of Pediatrics), Mexico City, Mexico,^35^School of Life Sciences, Ecole Polytechnique Fédérale de Lausanne, Lausanne, Switzerland; Precision Medicine Unit, Lausanne University Hospital and University of Lausanne, Lausanne, Switzerland, ^36^Research Unit, Hospital Universitario Nuestra Señora de Candelaria, Santa Cruz de Tenerife; CIBER de Enfermedades Respiratorias, Instituto de Salud Carlos III, Madrid; Genomics Division, Instituto Tecnológico y de Energías Renovables (ITER), Santa Cruz de Tenerife, Spain; Faculty of Health Sciences, University of Fernando Pessoa Canarias, Las Palmas de Gran Canaria, Spain, ^37^Group of Primary Immunodeficiencies, University of Antioquia UDEA, Medellin, Colombia, ^38^Pulmonology Department, Cliniques Universitaires Saint-Luc ; Institut de Recherche Expérimentale et Clinique (IREC), Université Catholique de Louvain, Brussels, Belgium, ^39^Sorbonne Université, Inserm, Centre d’Immunologie et des Maladies Infectieuses-Paris (CIMI PARIS), Assistance Publique-Hôpitaux de Paris (AP-HP) Hôpital Pitié-Salpêtrière, Paris, France, ^40^Feinstein Institute for Medical Research, Northwell Health USA, Manhasset, NY, USA, ^41^Center for Chronic Immunodeficiency & Institute for Immunodeficiency, Medical Center, Faculty of Medicine, University of Freiburg, Freiburg, Germany, ^42^Department of Paediatric Immunology and Pulmonology, Centre for Primary Immunodeficiency Ghent (CPIG), PID Research Laboratory, Jeffrey Modell Diagnosis and Research Centre, Ghent University Hospital, Ghent, Belgium, ^43^The Genetics Institute Tel Aviv Sourasky Medical Center, Tel Aviv, Israel, ^44^Sharjah Institute of Medical Research, College of Medicine, University of Sharjah, Sharjah, UAE, Prince Naif center for Immunology Research, King Saud University, Riyadh, SA, ^45^Division of Immunology, Department of Medical Biochemistry and Biophysics, Karolinska Institutet, Sweden, ^46^Institute for Systems Biology, Seattle, WA, USA, ^47^Departments of Pediatrics, Immunology and Microbiology, University of Colorado, School of Medicine, Aurora, CO, USA, ^48^Department of Medicine, Haukeland University Hospital, Bergen, Norway, ^49^Department of Community Pediatrics, Perinatal and Maternal Medicine, Tokyo Medical and Dental University (TMDU), Tokyo, Japan, ^50^Institute for Personalized Medicine, Icahn School of Medicine at Mount Sinai, New York, NY, USA; Department of Genetics and Genomic Sciences, Icahn School of Medicine at Mount Sinai, New York, NY, USA, ^51^Department of Medical Chemistry and Laboratory Medicine, Poznan University of Medical Sciences, Poznan, Poland, ^52^Bioinformatics and Applied Genomics Unit, Hellenic Pasteur Institute, Athens, Greece, ^53^Molecular Pathology, Department of Biomedicine, Institute of Biomedicine and Translational Medicine, University of Tartu, Tartu Estonia, ^54^Chang Gung University, Taoyuan County, Taiwan, ^55^Department of Paediatrics & Adolescent Medicine, The University of Hong Kong, Hong Kong, China, ^56^Shanghai Public Health Clinical Center, Fudan University, Shanghai, China, ^57^Department of Immunobiology, Yale University School of Medicine, New Haven, CT, USA, ^58^Zukerman Mind Brain Behavior Institute, Columbia University, New York, NY, USA, ^59^Department of Clinical Immunology and Infectious Diseases, National Research Institute of Tuberculosis and Lung Diseases, The Clinical Tuberculosis and Epidemiology Research Center, National Research Institute of Tuberculosis and Lung Diseases (NRITLD), Masih Daneshvari Hospital, Shahid Beheshti, University of Medical Sciences, Tehran, Iran, ^60^Primary Immunodeficiency Clinical Unit and Laboratory, Department of Dermatology, Venereology and Dermatooncology, Semmelweis University, Budapest, Hungary, ^61^Department of Pediatrics, University Hospitals Leuven; KU Leuven, Department of Microbiology, Immunology and Transplantation; Laboratory for Inborn Errors of Immunity, KU Leuven, Leuven, Belgium, ^62^Department of Pediatrics, Columbia University Irving Medical Center, New York, NY, USA, ^63^University Clinic for Children’s Diseases, Department of Pediatric Immunology, Medical Faculty, University “ StCyril and Methodij” Skopje, North Macedonia, ^64^Department of Biomedicine, Aarhus University, Aarhus, Denmark, ^65^Tokyo Medical & Dental University Hospital, Tokyo, Japan, ^66^A*STAR Infectious Disease Labs, Agency for Science, Technology and Research, Singapore; Lee Kong Chian School of Medicine, Nanyang Technology University, Singapore, ^67^National Institute of Allergy and Infectious Diseases, National Institutes of Health, Bethesda, MD, USA, ^68^Laboratory of Medical Genetics, IRCCS Bambino Gesù Children’s Hospital, Rome, Italy, ^69^Department of Biomedicine and Prevention, Tor Vergata University of Rome, Rome, Italy, ^70^Comparative Immunology Group, School of Biochemistry and Immunology, Trinity Biomedical Sciences Institute, Trinity College Dublin, Ireland, ^71^Department of Pediatrics, Graduate School of Biomedical and Health Sciences, Hiroshima University, Hiroshima, Japan, ^72^Tokyo Medical and Dental University, Tokyo, Japan, ^73^Department of Molecular Biology and Genetics, Bilkent University, Bilkent – Ankara, Turkey, ^74^Haitian Study Group for Kaposi’s Sarcoma and Opportunistic Infections (GHESKIO), Port-au-Prince, Haiti, ^75^Institute of Biomedical Research of IdiPAZ, University Hospital “La Paz”, Madrid, Spain, ^76^Institut de Biomedicina de València-CSIC, CIBERNED-ISCIII, Unitat Mixta de Neurologia i Genètica, IIS La Fe, Vallencia, Spain, ^77^Center for Discovery and Innovation, Hackensack Meridian Health, Nutley, NJ, USA, ^78^Department of Biosciences, Biotechnology and Environment, University of Bari A. Moro, Bari, Italy, ^79^IIBB-CSIC, IDIBAPS, Barcelona, Spain, ^80^Faculdades Pequeno Príncipe, Instituto de Pesquisa Pelé Pequeno Príncipe, Curitiba, Brazil, ^81^Neurometabolic Diseases Laboratory, Bellvitge Biomedical Research Institute (IDIBELL), L’Hospitalet de Llobregat, Barcelona, Spain; Catalan Institution of Research and Advanced Studies (ICREA), Barcelona, Spain; Center for Biomedical Research on Rare Diseases (CIBERER), ISCIII, Barcelona, Spain, ^82^Human Evolutionary Genetics Unit, CNRS U2000, Institut Pasteur, Paris, France; Human Genomics and Evolution, Collège de France, Paris, France, ^83^Al Jalila Children’s Hospital, Dubai, UAE, ^84^University Hospital St Marina, Varna, Bulgaria, ^85^Department of Immunology, University Hospital of Gran Canaria Dr Negrín, Canarian Health System, Las Palmas de Gran Canaria; Department of Clinical Sciences, University Fernando Pessoa Canarias, Las Palmas de Gran Canaria, Spain; Department of Medical and Surgical Sciences, School of Medicine, University of Las Palmas de Gran Canaria, Las Palmas de Gran Canaria, Spain, ^86^Department of Paediatric Infectious Diseases and Virology, Imperial College London, London, UK; Centre for Paediatrics and Child Health, Faculty of Medicine, Imperial College London, London, UK, ^87^Department of Immunology, Second Faculty of Medicine Charles University, V Uvalu, University Hospital in Motol, Prague, Czech Republic, ^88^Adult Immunodeficiency Unit, Infectious Diseases, Inflammation Center, University of Helsinki and Helsinki University Hospital, Helsinki, Finland; Rare Diseases Center and Pediatric Research Center, Children’s Hospital, University of Helsinki and Helsinki University Hospital, Helsinki, Finland, ^89^Dr. Shahrooei Lab, 22 Bahman St., Ashrafi Esfahani Blvd, Tehran, Iran; Clinical and Diagnostic Immunology lab, Department of Microbiology, Immunology, and Transplantation, KU Leuven, Leuven, Belgium, ^90^Department of Immunology, Dmitry Rogachev National Medical Research Center of Pediatric Hematology, Oncology and Immunology, Moscow, Russia, ^91^Central European Institute of Technology & Department of Biology, Faculty of Medicine, Masaryk University, Brno, Czech Republic, ^92^Department of Pharmacology & Molecular Therapeutics, Uniformed Services University of the Health Sciences, Bethesda, MD, USA, ^93^Pediatric Infectious Diseases and Immunodeficiencies Unit, Hospital Universitari Vall d’Hebron, Vall d’Hebron Research Institute, Vall d’Hebron Barcelona Hospital Campus, Universitat Autònoma de Barcelona (UAB), Barcelona, Catalonia, Spain, ^94^Université de Paris, Institut de Recherche Saint-Louis, INSERM U976, Hôpital Saint-Louis, Paris, France; AP-HP, Hôpital Saint-Louis, Laboratoire d’Immunologie, Paris, France, ^95^St Giles Laboratory of Human Genetics of Infectious Diseases, Rockefeller Branch, The Rockefeller University, New York, NY, USA; Department of Medical Microbiology, University Medical Center Utrecht, Utrecht University, Utrecht, Netherlands, ^96^Department of Internal Medicine II, Medical University of Innsbruck, Innsbruck, Austria, ^97^Garvan Institute of Medical Research, Darlinghurst, NSW, Australia; St Vincent’s Clinical School, Faculty of Medicine, UNSW Sydney, NSW, Australia, ^98^Departments of Medical Genetics & Histology and Embryology, Faculty of Medicine; Department of Translational Medicine, Health Sciences Institude, Bursa Uludağ University, Bursa, Turkey, ^99^Precision Medicine Unit, Lausanne University Hospital and University of Lausanne, Lausanne, Switzerland, ^100^Etablissement Francais Du Sang, La Plaine-Saint Denis, Saint-Denis, France, ^101^Hospices Civils de Lyon, Lyon, France; International Center of Research in Infectiology, Lyon University, INSERM U1111, CNRS UMR 5308, ENS, UCBL, Lyon, France, ^102^BC Children’s Hospital, The University of British Columbia, Vancouver, Canada, ^103^Centre for Precision Therapeutics, Genetics & Genomic Medicine Centre, NeuroGen Children’s Healthcare and Lecturer, Holy Family Red Crescent Medical College Dhaka, Bangladesh, ^104^College of Medicine, Mohammed Bin Rashid University of Medicine and Health Sciences, Dubai, UAE; Cellular Intelligence (Ci) Lab, GenomeArc Inc, Toronto, ON, Canada, ^105^Department of Neurology, Amsterdam Neuroscience, Amsterdam University Medical Center, University of Amsterdam, Amsterdam, The Netherlands, ^106^Department of Medicine, Division of Infectious Diseases, McGill University Health Centre, Montréal, Québec, Canada; Infectious Disease Susceptibility Program, Research Institute, McGill University Health Centre, Montréal, Québec, Canada, ^107^Department of Pediatric Pneumology, Immunology and Intensive Care, Charité Universitätsmedizin, Berlin University Hospital Center, Berlin, Germany; Labor Berlin GmbH, Department of Immunology, Berlin, Germany; Berlin Institutes of Health (BIH), Berlin-Brandenburg Center for Regenerative Therapies, Berlin, Germany, ^108^Department of General Internal Medicine, Medical Intensive Care Unit, University Hospitals Leuven, Leuven, Belgium, ^109^Biosciences Institute, University of São Paulo, São Paulo, Brazil, ^110^Molecular Biophysics Division, Faculty of Physics, A Mickiewicz University, Poznań, Poland.

***Members of COVID-STORM Clinicians*:** Giuseppe Foti^1^, Giuseppe Citerio^1^, Ernesto Contro^1^, Alberto Pesci^2^, Maria Grazia Valsecchi^3^, Marina Cazzaniga^4^, Giacomo Bellani^5^.

^1^Department of Emergency, Anesthesia and Intensive Care, School of Medicine and Surgery, University of Milano-Bicocca, San Gerardo Hospital, Monza, Italy. ^2^Department of Pneumology, School of Medicine and Surgery, University of Milano-Bicocca, San Gerardo Hospital, Monza, Italy. ^3^Center of Bioinformatics and Biostatistics, School of Medicine and Surgery, University of Milano-Bicocca, San Gerardo Hospital, Monza, Italy. ^4^Phase I Research Center, School of Medicine and Surgery, University of Milano-Bicocca, San Gerardo Hospital, Monza, Italy. ^5^Interdepartmental Centre for Medical Sciences (CISMed), University of Trento, Trento, Italy.

***Members of COVID Clinicians*:** Jorge Abad^1^, Giulia Accordino^2^, Micol Angelini^3^, Sergio Aguilera-Albesa^4^, Aina Aguiló-Cucurull^5^, Alessandro Aiuti^6^, Esra Akyüz Özkan^7^, Ilad Alavi Darazam^8^, Jonathan Antonio Roblero Albisures^9^, Juan C. Aldave^10^, Miquel Alfonso Ramos^11^, Taj Ali Khan^12^, Anna Aliberti^13^, Seyed Alireza Nadji^14^, Gulsum Alkan^15^, Suzan A. AlKhater^16^, Jerome Allardet-Servent^17^, Luis M. Allende^18^, Rebeca Alonso-Arias^19^, Mohammed S. Alshahrani^20^, Laia Alsina^21^, Marie-Alexandra Alyanakian^22^, Blanca Amador Borrero^23^, Zahir Amoura^24^, Arnau Antolí^25^, Romain Arrestier^26^, Mélodie Aubart^27^, Teresa Auguet^28^, Iryna Avramenko^29^, Gökhan Aytekin^30^, Axelle Azot^31^, Seiamak Bahram^32^, Fanny Bajolle^33^, Fausto Baldanti^34^, Aurélie Baldolli^35^, Maite Ballester^36^, Hagit Baris Feldman^37^, Benoit Barrou^38^, Federica Barzaghi^6^, Sabrina Basso^39^, Gulsum Iclal Bayhan^40^, Alexandre Belot^41^, Liliana Bezrodnik^42^, Agurtzane Bilbao^43^, Geraldine Blanchard-Rohner^44^, Ignacio Blanco^45^, Adeline Blandinières^46^, Daniel Blázquez-Gamero^47^, Alexandre Bleibtreu^48^, Marketa Bloomfield^49^, Mireia Bolivar-Prados^50^, Anastasiia Bondarenko^51^, Alessandro Borghesi^3^, Raphael Borie^52^, Elisabeth Botdhlo-Nevers^53^, Ahmed A. Bousfiha^54^, Aurore Bousquet^55^, David Boutolleau^56^, Claire Bouvattier^57^, Oksana Boyarchuk^58^, Juliette Bravais^59^, M. Luisa Briones^60^, Marie-Eve Brunner^61^, Raffaele Bruno^62^, Maria Rita P. Bueno^63^, Huda Bukhari^64^, Jacinta Bustamante^33^, Juan José Cáceres Agra^65^, Ruggero Capra^66^, Raphael Carapito^67^, Maria Carrabba^68^, Giorgio Casari^6^, Carlos Casasnovas^69^, Marion Caseris^70^, Irene Cassaniti^34^, Martin Castelle^71^, Francesco Castelli^72^, Martín Castillo de Vera^73^, Mateus V. Castro^63^, Emilie Catherinot^74^, Jale Bengi Celik^75^, Alessandro Ceschi^76^, Martin Chalumeau^77^, Bruno Charbit^78^, Cécile Boulanger^79^, Père Clavé^50^, Bonaventura Clotet^80^, Anna Codina^81^, Yves Cohen^82^, Roger Colobran^83^, Cloé Comarmond^84^, Alain Combes^85^, Patrizia Comoli^39^, Angelo G. Corsico^2^, Taner Coşkuner^86^, Aleksandar Cvetkovski^87^, Cyril Cyrus^88^, David Dalmau^89^, François Danion^90^, David Ross Darley^91^, Vincent Das^92^, Nicolas Dauby^93^, Stéphane Dauger^94^, Paul De Munter^95^, Loic de Pontual^96^, Amin Dehban^97^, Geoffroy Delplancq^98^, Alexandre Demoule^99^, Isabelle Desguerre^100^, Antonio Di Sabatino^101^, Jean-Luc Diehl^102^, Stephanie Dobbelaere^103^, Elena Domínguez-Garrido^104^, Clément Dubost^105^, Olov Ekwall^106^, Şefika Elmas Bozdemir^107^, Marwa H. Elnagdy^108^, Melike Emiroglu^15^, Akifumi Endo^109^, Emine Hafize Erdeniz^110^, Selma Erol Aytekin^111^, Maria Pilar Etxart Lasa^112^, Romain Euvrard^113^, Giovanna Fabio^68^, Laurence Faivre^114^, Antonin Falck^115^, Muriel Fartoukh^116^, Morgane Faure^117^, Miguel Fernandez Arquero^118^, Ricard Ferrer^119^, Jose Ferreres^120^, Carlos Flores^121^, Bruno Francois^122^, Victoria Fumadó^123^, Kitty S. C. Fung^124^, Francesca Fusco^125^, Alenka Gagro^126^, Blanca Garcia Solis^127^, Pierre Garçon^343^, Pascale Gaussem^128^, Zeynep Gayretli^129^, Juana Gil-Herrera^130^, Laurent Gilardin^131^, Audrey Giraud Gatineau^132^, Mònica Girona-Alarcón^133^, Karen Alejandra Cifuentes Godínez^134^, Jean-Christophe Goffard^135^, Nacho Gonzales^136^, Luis I. Gonzalez-Granado^137^, Rafaela González-Montelongo^138^, Antoine Guerder^139^, Belgin Gülhan^140^, Victor Daniel Gumucio^141^, Leif Gunnar Hanitsch^142^, Jan Gunst^143^, Marta Gut^144^, Jérôme Hadjadj^145^, Filomeen Haerynck^146^, Rabih Halwani^147^, Lennart Hammarström^148^, Selda Hancerli^149^, Tetyana Hariyan^150^, Nevin Hatipoglu^151^, Deniz Heppekcan^152^, Elisa Hernandez-Brito^153^, Po-ki Ho^154^, María Soledad Holanda-Peña^155^, Juan P. Horcajada^156^, Sami Hraiech^157^, Linda Humbert^158^, Ivan F. N. Hung^159^, Alejandro D. Iglesias^160^, Antonio Íñigo-Campos^138^, Matthieu Jamme^161^, María Jesús Arranz^89^, Marie-Thérèse Jimeno^162^, Iolanda Jordan^133^, Saliha Kanık-Yüksek^163^, Yalcin Kara^164^, Aydın Karahan^165^, Adem Karbuz^166^, Kadriye Kart Yasar^167^, Ozgur Kasapcopur^168^, Kenichi Kashimada^169^, Sevgi Keles^111^, Yasemin Kendir Demirkol^170^, Yasutoshi Kido^171^, Can Kizil^172^, Ahmet Osman Kılıç^173^, Adam Klocperk^174^, Antonia Koutsoukou^175^, Zbigniew J. Król^176^, Hatem Ksouri^177^, Paul Kuentz^178^, Arthur M. C. Kwan^179^, Yat Wah M. Kwan^180^, Janette S. Y. Kwok^181^, Jean-Christophe Lagier^182^, David S. Y. Lam^183^, Vicky Lampropoulou^184^, Fanny Lanternier^185^, Yu-Lung Lau^186^, Fleur Le Bourgeois^94^, Yee-Sin Leo^187^, Rafael Leon Lopez^188^, Daniel Leung^186^, Michael Levin^189^, Michael Levy^94^, Romain Lévy^33^, Zhi Li^78^, Daniele Lilleri^34^, Edson Jose Adrian Bolanos Lima^190^, Agnes Linglart^191^, Eduardo López-Collazo^192^, José M. Lorenzo-Salazar^138^, Céline Louapre^193^, Catherine Lubetzki^193^, Kwok-Cheung Lung^194^, Charles-Edouard Luyt^195^, David C. Lye^196^, Cinthia Magnone^197^, Davood Mansouri^198^, Enrico Marchioni^199^, Carola Marioli^2^, Majid Marjani^200^, Laura Marques^201^, Jesus Marquez Pereira^202^, Andrea Martín-Nalda^203^, David Martínez Pueyo^204^, Javier Martinez-Picado^205^, Iciar Marzana^206^, Carmen Mata-Martínez^207^, Alexis Mathian^24^, Larissa R. B. Matos^63^, Gail V. Matthews^208^, Julien Mayaux^209^, Raquel McLaughlin-Garcia^210^, Philippe Meersseman^211^, Jean-Louis Mège^212^, Armand Mekontso-Dessap^213^, Isabelle Melki^115^, Federica Meloni^346^, Jean-François Meritet^214^, Paolo Merlani^215^, Özge Metin Akcan^216^, Isabelle Meyts^217^, Mehdi Mezidi^218^, Isabelle Migeotte^219^, Maude Millereux^220^, Matthieu Million^221^, Tristan Mirault^222^, Clotilde Mircher^223^, Mehdi Mirsaeidi^224^, Yoko Mizoguchi^225^, Bhavi P. Modi^226^, Francesco Mojoli^13^, Elsa Moncomble^227^, Abián Montesdeoca Melián^228^, Antonio Morales Martinez^229^, Francisco Morandeira^230^, Pierre-Emmanuel Morange^231^, Clémence Mordacq^158^, Guillaume Morelle^232^, Stéphane J. Mouly^233^, Adrián Muñoz-Barrera^138^, Cyril Nafati^234^, Shintaro Nagashima^235^, Yu Nakagama^171^, Bénédicte Neven^236^, João Farela Neves^237^, Lisa F. P. Ng^238^, Yuk-Yung Ng^239^, hubert Nielly^105^, Yeray Novoa Medina^210^, Esmeralda Nuñez Cuadros^240^, Semsi Nur Karabela^167^, J. Gonzalo Ocejo-Vinyals^241^, Keisuke Okamoto^109^, Mehdi Oualha^33^, Amani Ouedrani^22^, Tayfun Özçelik^242^, Aslinur Ozkaya-Parlakay^140^, Michele Pagani^13^, Qiang Pan-Hammarström^148^, Maria Papadaki^243^, Christophe Parizot^209^, Philippe Parola^244^, Tiffany Pascreau^245^, Stéphane Paul^246^, Estela Paz-Artal^247^, Sigifredo Pedraza^248^, Nancy Carolina González Pellecer^134^, Silvia Pellegrini^249^, Rebeca Pérez de Diego^127^, Xosé Luis Pérez-Fernández^141^, Aurélien Philippe^250^, Quentin Philippot^116^, Adrien Picod^251^, Marc Pineton de Chambrun^85^, Antonio Piralla^34^, Laura Planas-Serra^252^, Dominique Ploin^253^, Julien Poissy^254^, Géraldine Poncelet^70^, Garyphallia Poulakou^175^, Marie S. Pouletty^255^, Persia Pourshahnazari^256^, Jia Li Qiu-Chen^257^, Paul Quentric^209^, Thomas Rambaud^258^, Didier Raoult^212^, Violette Raoult^259^, Anne-Sophie Rebillat^223^, Claire Redin^260^, Léa Resmini^261^, Pilar Ricart^262^, Jean-Christophe Richard^263^, Raúl Rigo-Bonnin^264^, Nadia rivet^46^, Jacques G. Rivière^265^, Gemma Rocamora-Blanch^25^, Mathieu P. Rodero^266^, Carlos Rodrigo^267^, Luis Antonio Rodriguez^190^, Carlos Rodriguez-Gallego^268^, Agustí Rodriguez-Palmero^269^, Carolina Soledad Romero^270^, Anya Rothenbuhler^271^, Damien Roux^272^, Nikoletta Rovina^175^, Flore Rozenberg^273^, Yvon Ruch^90^, Montse Ruiz^274^, Maria Yolanda Ruiz del Prado^275^, Juan Carlos Ruiz-Rodriguez^119^, Joan Sabater-Riera^141^, Kai Saks^276^, Maria Salagianni^184^, Oliver Sanchez^277^, Adrián Sánchez-Montalvá^278^, Silvia Sánchez-Ramón^279^, Laire Schidlowski^280^, Agatha Schluter^252^, Julien Schmidt^281^, Matthieu Schmidt^282^, Catharina Schuetz^283^, Cyril E. Schweitzer^284^, Francesco Scolari^285^, Anna Sediva^286^, Luis Seijo^287^, Analia Gisela Seminario^42^, Damien Sene^23^, Piseth Seng^221^, Sevtap Senoglu^167^, Mikko Seppänen^288^, Alex Serra Llovich^289^, Mohammad Shahrooei^97^, Anna Shcherbina^290^, Virginie Siguret^291^, Eleni Siouti^292^, David M. Smadja^293^, Nikaia Smith^78^, Ali Sobh^294^, Xavier Solanich^25^, Jordi Solé-Violán^295^, Catherine Soler^296^, Pere Soler-Palacín^297^, Betül Sözeri^86^, Giulia Maria Stella^2^, Yuriy Stepanovskiy^298^, Annabelle Stoclin^299^, Fabio Taccone^219^, Yacine Tandjaoui-Lambiotte^300^, Jean-Luc Taupin^301^, Simon J. Tavernier^302^, Loreto Vidaur Tello^112^, Benjamin Terrier^303^, Guillaume Thiery^304^, Christian Thorball^260^, Karolina Thorn^305^, Caroline Thumerelle^158^, Imran Tipu^306^, Martin Tolstrup^307^, Gabriele Tomasoni^308^, Julie Toubiana^77^, Josep Trenado Alvarez^309^, Vasiliki Triantafyllia^310^, Sophie Trouillet-Assant^311^, Jesús Troya^312^, Owen T. Y. Tsang^313^, Liina Tserel^314^, Eugene Y. K. Tso^315^, Alessandra Tucci^316^, Şadiye Kübra Tüter Öz^15^, Matilde Valeria Ursini^125^, Takanori Utsumi^225^, Yurdagul Uzunhan^317^, Pierre Vabres^318^, Juan Valencia-Ramos^319^, Ana Maria Van Den Rym^127^, Isabelle Vandernoot^320^, Valentina Velez-Santamaria^321^, Silvia Patricia Zuniga Veliz^134^, Mateus C. Vidigal^322^, Sébastien Viel^253^, Cédric Villain^323^, Marie E. Vilaire-Meunier^223^, Judit Villar-García^324^, Audrey Vincent^57^, Dimitri Van der Linden^325^, Guillaume Voiriot^326^, Alla Volokha^327^, Fanny Vuotto^158^, Els Wauters^328^, Joost Wauters^329^, Alan K. L. Wu^330^, Tak-Chiu Wu^331^, Aysun Yahşi^332^, Osman Yesilbas^333^, Mehmet Yildiz^168^, Barnaby E. Young^187^, Ufuk Yükselmiş^334^, Mayana Zatz^63^, Marco Zecca^39^, Valentina Zuccaro^62^, Jens Van Praet^335^, Bart N. Lambrecht^336^, Eva Van Braeckel^336^, Cédric Bosteels^336^, Levi Hoste^337^, Eric Hoste^338^, Fré Bauters^336^, Jozefien De Clercq^336^, Catherine Heijmans^339^, Hans Slabbynck^340^, Leslie Naesens^341^, Benoit Florkin^342^, Mary-Anne Young^344^, Amanda Willis^344^, Paloma Lapuente-Suanzes^345^, Ana de Andrés-Martín^345^.

^1^Germans Trias i Pujol University Hospital and Research Institute, Badalona, Barcelona, Spain. ^2^Respiratory Diseases Division, IRCCS Policlinico San Matteo Foundation, University of Pavia, Pavia, Italy. ^3^Neonatal Intensive Care Unit, Fondazione IRCCS Policlinico San Matteo, Pavia, Italy. ^4^Navarra Health Service Hospital, Pamplona, Spain. ^5^Jeffrey Modell Diagnostic and Research Center for Primary Immunodeficiencies, Barcelona, Catalonia, Spain; Immunology Division, Genetics Department, Vall d’Hebron University Hospital (HUVH), Vall d’Hebron Research Institute (VHIR), Vall d’Hebron Barcelona Hospital Campus, Universitat Autònoma de Barcelona (UAB), Barcelona, Catalonia, Spain. ^6^Immunohematology Unit, San Raffaele Hospital, Milan, Italy. ^7^Ondokuz Mayıs University Medical Faculty Pediatrics, Samsun, Turkey. ^8^Department of Infectious Diseases, Loghman Hakim Hospital, Shahid Beheshti University of Medical Sciences, Tehran, Iran. ^9^Hospital Regional de Huehuetenango, “Dr. Jorge Vides de Molina,” Huehuetenango, Guatemala. ^10^Hospital Nacional Edgardo Rebagliati Martins, Lima, Peru. ^11^Parc Sanitari Sant Joan de Déu, Sant Boi de Llobregat Spain. ^12^Khyber Medical University, Khyber Pakhtunkhwa, Pakistan. ^13^Anesthesia and Intensive Care, Rianimazione I, Fondazione IRCCS Policlinico San Matteo, Pavia, Italy. ^14^Virology Research Center, National Institutes of Tuberculosis and Lung Diseases, Shahid Beheshti University of Medical Sciences, Tehran, Iran. ^15^Department of Pediatrics, Division of Pediatric Infectious Diseases, Selcuk University Faculty of Medicine, Konya, Turkey. ^16^College of Medicine, Imam Abdulrahman Bin Faisal University, Dammam, Saudi Arabia; Department of Pediatrics, King Fahad Hospital of the University, Al-Khobar, Saudi Arabia. ^17^Intensive Care Unit, Hôpital Européen, Marseille, France. ^18^Immunology Department, Hospital 12 de Octubre, Research Institute imas12, Complutense University, Madrid, Spain. ^19^Immunology Department, Asturias Central University Hospital, Biosanitary Research Institute of the Principality of Asturias (ISPA), Oviedo, Spain. ^20^Emergency and Critical Care Medicine Departments, College of Medicine, Imam AbdulRahman Ben Faisal University, Dammam, Saudi Arabia. ^21^Clinical Immunology and Primary Immunodeficiencias Unit, Hospital Sant Joan de Déu, Institut de Recerca Sant Joan de Déu, Barcelona, Spain; Universitat de Barcelona, Barcelona, Spain. ^22^Department of Biological Immunology, Necker Hospital for Sick Children, AP-HP and INEM, Paris, France. ^23^Internal Medicine Department, Hôpital Lariboisière, AP-HP, Paris, France; Université de Paris, Paris, France. ^24^Internal Medicine Department, Pitié-Salpétrière Hospital, Paris, France. ^25^Department of Internal Medicine, Hospital Universitari de Bellvitge, IDIBELL, Barcelona, Spain. ^26^Service de Médecine Intensive Réanimation, Hôpitaux Universitaires Henri Mondor, AP-HP, Créteil, France; Groupe de Recherche Clinique CARMAS, Faculté de Santé de Créteil, Université Paris Est Créteil, Créteil, France. ^27^INSERM U1163, University of Paris, Imagine Institute, Paris, France and Pediatric Neurology Department, Necker-Enfants malades Hospital, AP-HP, Paris, France. ^28^Hospital U. de Tarragona Joan XXIII. Universitat Rovira i Virgili (URV). IISPV, Tarragona, Spain. ^29^Department of Propedeutics of Pediatrics and Medical Genetics, Danylo Halytsky Lviv National Medical University, Lviv, Ukraine. ^30^Department of Immunology and Allergy, Konya City Hospital, Konya, Turkey. ^31^Private Practice, Paris, France. ^32^INSERM U1109, University of Strasbourg, Strasbourg, France. ^33^Necker Hospital for Sick Children, AP-HP, Paris, France. ^34^Molecular Virology Unit, Microbiology and Virology Department, Fondazione IRCCS Policlinico San Matteo, Pavia, Italy. ^35^Department of Infectious Diseases, CHU de Caen, Caen, France. ^36^Consorcio Hospital General Universitario, Valencia, Spain. ^37^Genetics Institute, Tel Aviv Sourasky Medical Center and Sackler Faculty of Medicine, Tel Aviv University, Tel Aviv, Israel. ^38^Department of Urology, Nephrology, Transplantation, APHP-SU, Sorbonne Université, INSERM U 1082, Paris, France. ^39^Cell Factory and Pediatric Hematology-Oncology, Fondazione IRCCS Policlinico San Matteo, Pavia, Italy. ^40^Yildirim Beyazit University, Faculty of Medicine, Ankara City Hospital, Children’s Hospital, Ankara, Turkey. ^41^University of Lyon, CIRI, INSERM U1111, National Referee Centre RAISE, Pediatric Rheumatology, HFME, Hospices Civils de Lyon, Lyon, France. ^42^Center for Clinical Immunology, CABA, Buenos Aires, Argentina. ^43^Cruces University Hospital, Bizkaia, Spain. ^44^Paediatric Immunology and Vaccinology Unit, Geneva University Hospitals and Faculty of Medicine, Geneva, Switzerland. ^45^University Hospital and Research Institute “Germans Trias i Pujol,” Badalona, Spain. ^46^Hematology, Georges Pompidou Hospital, AP-HP, Paris, France. ^47^Pediatric Infectious Diseases Unit, Instituto de Investigación Hospital 12 de Octubre (imas12), Hospital Universitario 12 de Octubre, Universidad Complutense, Madrid, Spain. ^48^Infectious disease Unit, Pitié-Salpêtrière Hospital, AP-AP, Paris, France. ^49^Department of Pediatrics, Thomayer’s Hospital, first Faculty of Medicine, Charles University, Prague, Czech Republic; Department of Immunology, Motol University Hospital, Second Faculty of Medicine, Charles University, Prague, Czech Republic. ^50^Centro de Investigación Biomédica en Red de Enfermedades Hepàticas y Digestivas (Ciberehd), Hospital de Mataró, Consorci Sanitari del Maresme, Mataró, Spain. ^51^Shupyk National Healthcare University of Ukraine, Kyiv, Ukraine. ^52^Service de Pneumologie, Hopital Bichat, AP-HP, Paris, France. ^53^Department of Infectious Diseases, CIC1408, GIMAP CIRI INSERM U1111, University Hospital of Saint-Etienne, Saint-Etienne, France. ^54^Clinical Immunology Unit, Pediatric Infectious Disease Department, Faculty of Medicine and Pharmacy, Averroes University Hospital, LICIA Laboratoire d’immunologie clinique, d’inflammation et d’allergie, Hassann Ii University, Casablanca, Morocco. ^55^Bégin Military Hospital, St Mandé, France. ^56^Sorbonne Université, INSERM, Institut Pierre Louis d’Epidémiologie et de Santé Publique (iPLESP), AP-HP, Hôpital Pitié Salpêtrière, Service de Virologie, Paris, France. ^57^Endocrinology Unit, AP-HP Hôpitaux Universitaires Paris-Sud, Le Kremlin-Bicêtre, France. ^58^Department of Children’s Diseases and Pediatric Surgery, I. Horbachevsky Ternopil National Medical University, Ternopil, Ukraine. ^59^Pneumology Unit, Tenon Hospital, AP-HP, Paris, France. ^60^Department of Respiratory Diseases, Hospital Clínico y Universitario de Valencia, Valencia, Spain. ^61^Intensive Care Unit, Réseau Hospitalier Neuchâtelois, Neuchâtel, Switzerland. ^62^Infectious Diseases Unit, Fondazione IRCCS Policlinico San Matteo, Pavia, Italy. ^63^Human Genome and Stem Cell Research Center, University of São Paulo, São Paulo, Brazil. ^64^Department of Internal Medicine, College of Medicine, Imam Abdulrahman Bin Faisal University, Dammam, Saudi Arabia. ^65^Hospital Insular, Las Palmas de Gran Canaria, Spain. ^66^MS Center, Spedali Civili, Brescia, Italy. ^67^Laboratoire d’ImmunoRhumatologie Moléculaire, plateforme GENOMAX, INSERM UMR_S 1109, Faculté de Médecine, ITI TRANSPLANTEX NG, Université de Strasbourg, Strasbourg, France. ^68^Fondazione IRCCS Ca′ Granda Ospedale Maggiore Policlinico, Milan, Italy. ^69^Neuromuscular Unit, Neurology Department, Hospital Universitari de Bellvitge–IDIBELL and CIBERER, Barcelona, Spain. ^70^Hopital Robert Debré, Paris, France. ^71^Pediatric Immunohematology Unit, Necker Enfants Malades Hospital, AP-HP, Paris, France. ^72^Department of Infectious and Tropical Diseases, University of Brescia, ASST Spedali Civili di Brescia, Brescia, Italy. ^73^Doctoral Health Care Center, Canarian Health System, Las Palmas de Gran Canaria, Spain. ^74^Hôpital Foch, Suresnes, France. ^75^Selcuk University Faculty of Medicine, Department of Anesthesiology and Reanimation, Intensive Care Medicine Unit, Konya, Turkey. ^76^Division of Clinical Pharmacology and Toxicology, Institute of Pharmacological Sciences of Southern Switzerland, Ente Ospedaliero Cantonale and Faculty of Biomedical Sciences, Università della Svizzera italiana, Lugano, Switzerland. ^77^Necker Hospital for Sick Children, Paris University, AP-HP, Paris, France. ^78^Pasteur Institute, Paris, France. ^79^Department of Pediatric Hemato-oncology, UCL Louvain, Brussels, Belgium. ^80^University Hospital and Research Institute “Germans Trias i Pujol,” IrsiCaixa AIDS Research Institute, UVic-UCC, Badalona, Spain. ^81^Clinical Biochemistry, Pathology, Paediatric Neurology and Molecular Medicine Departments and Biobank, Institut de Recerca Sant Joan de Déu and CIBERER-ISCIII, Esplugues, Spain. ^82^AP-HP, Avicenne Hospital, Intensive Care Unit, Bobigny, France; University Sorbonne Paris Nord, Bobigny, France; INSERM, U942, F-75010, Paris, France. ^83^Hospital Universitari Vall d’Hebron, Barcelona, Spain. ^84^Pitié-Salpêtrière Hospital, Paris, France. ^85^Service de médecine Intensive Réanimation, Groupe Hospitalier Pitié-Salpêtrière, Sorbonne Université, Paris, France. ^86^Umraniye Training and Research Hospital, Istanbul, Turkey. ^87^Faculty of Medical Sciences at University “Goce Delcev,” Shtip, North Macedonia. ^88^Department of Biochemistry, College of Medicine, Imam Abdulrahman Bin Faisal University, Dammam, Saudi Arabia. ^89^Fundació Docencia i Recerca Mutua Terrassa, Barcelona, Spain. ^90^Maladies Infectieuses et Tropicales, Nouvel Hôpital Civil, CHU Strasbourg, Strasbourg, France. ^91^UNSW Medicine, St Vincent’s Clinical School, Sydney, NSW, Australia; Department of Thoracic Medicine, St Vincent’s Hospital Darlinghurst, Sydney, NSW, Australia. ^92^Intensive Care Unit, Montreuil Hospital, Montreuil, France. ^93^CHU Saint-Pierre, Université Libre de Bruxelles (ULB), Brussels, Belgium. ^94^Pediatric Intensive Care Unit, Robert-Debré University Hospital, AP-HP, Paris, France. ^95^General Internal Medicine, University Hospitals Leuven, Leuven, Belgium. ^96^Hôpital Jean Verdier, AP-HP, Bondy, France. ^97^Dr. Shahrooei Lab, 22 Bahman St., Ashrafi Esfahani Blvd, Tehran, Iran; Clinical and Diagnostic Immunology lab, Department of Microbiology, Immunology, and Transplantation, KU Leuven, Leuven, Belgium. ^98^Centre de génétique humaine, CHU Besançon, Besançon, France. ^99^Sorbonne Université médecine and AP-HP Sorbonne université site Pitié-Salpêtrière, Paris, France. ^100^Pediatric Neurology Department, Necker-Enfants Malades Hospital, AP-HP, Paris, France. ^101^Department of Internal Medicine, Fondazione IRCCS Policlinico San Matteo, University of Pavia, Pavia, Italy. ^102^Intensive Care Unit, Georges Pompidou Hospital, AP-HP, Paris, France. ^103^Department of Pneumology, AZ Delta, Roeselare, Belgium. ^104^Molecular Diagnostic Unit, Fundación Rioja Salud, Logroño, La Rioja, Spain. ^105^Bégin Military Hospital, Saint Mandé, France. ^106^Department of Pediatrics, Institute of Clinical Sciences, Sahlgrenska Academy, University of Gothenburg, Gothenburg, Sweden; Department of Rheumatology and Inflammation Research, Institute of Medicine, Sahlgrenska Academy, University of Gothenburg, Gothenburg, Sweden. ^107^Bursa City Hospital, Bursa, Turkey. ^108^Department of Medical Biochemistry and Molecular Biology, Faculty of Medicine, Mansoura University, Mansoura, Egypt. ^109^Tokyo Medical and Dental University, Tokyo, Japan. ^110^Ondokuz Mayıs University Faculty of Medicine, Samsun, Turkey. ^111^Necmettin Erbakan University, Meram Medical Faculty, Division of Pediatric Allergy and Immunology, Konya, Turkey. ^112^Intensive Care Medicine, Donostia University Hospital, Biodonostia Institute of Donostia, CIBER Enfermedades Respiratorias ISCIII, Donostia, Spain. ^113^Internal Medicine, University Hospital Edouard Herriot, Hospices Civils de Lyon, Lyon, France. ^114^Centre de Génétique, CHU Dijon, Dijon, France. ^115^Robert Debré Hospital, Paris, France. ^116^AP-HP Tenon Hospital, Paris, France. ^117^Sorbonne Universités, UPMC University of Paris, Paris, France. ^118^Department of Clinical Immunology, Hospital Clínico San Carlos, Madrid, Spain. ^119^Intensive Care Department, Vall d’Hebron University Hospital (HUVH), Vall d’Hebron Barcelona Hospital Campus, Barcelona, Catalonia, Spain; Shock, Organ Dysfunction and Resuscitation Research Group, Vall d’Hebron Research Institute (VHIR), Vall d’Hebron Barcelona Hospital Campus, Barcelona, Catalonia, Spain. ^120^Intensive Care Unit, Hospital Clínico y Universitario de Valencia, Valencia, Spain. ^121^Genomics Division, Instituto Tecnológico y de Energías Renovables (ITER), Santa Cruz de Tenerife, Spain; CIBER de Enfermedades Respiratorias, Instituto de Salud Carlos III, Madrid, Spain; Research Unit, Hospital Universitario N.S. de Candelaria, Santa Cruz de Tenerife, Spain; Faculty of Health Sciences, University of Fernando Pessoa Canarias, Las Palmas de Gran Canaria, Spain. ^122^CHU Limoges and INSERM CIC 1435 and UMR 1092, Limoges, France. ^123^Infectious Diseases Unit, Department of Pediatrics, Hospital Sant Joan de Déu, Barcelona, Spain; Institut de Recerca Sant Joan de Déu, Spain; Universitat de Barcelona (UB), Barcelona, Spain. ^124^Department of Pathology, United Christian Hospital, Hong Kong, China. ^125^Institute of Genetics and Biophysics “Adriano Buzzati-Traverso,” IGB-CNR, Naples, Italy. ^126^Department of Pediatrics, Children’s Hospital Zagreb, University of Zagreb School of Medicine, Zagreb, Josip Juraj Strossmayer University of Osijek, Medical Faculty Osijek, Osijek, Croatia. ^127^Laboratory of Immunogenetics of Human Diseases, IdiPAZ Institute for Health Research, La Paz Hospital, Madrid, Spain. ^128^Hematology, AP-HP, Hopital Européen Georges Pompidou and INSERM UMR-S1140, Paris, France. ^129^Faculty of Medicine, Department of Pediatrics, Division of Pediatric Infectious Diseases, Karadeniz Technical University, Trabzon, Turkey. ^130^Division of Immunology, Hospital General Universitario and Instituto de Investigación Sanitaria “Gregorio Marañón,” Madrid, Spain. ^131^Bégin Military Hospital, Bégin, France. ^132^Aix Marseille Univ, IRD, AP-HM, SSA, VITROME, IHU Méditerranée Infection, Marseille, France, French Armed Forces Center for Epidemiology and Public Health (CESPA), Marseille, France. ^133^Pediatric Intensive Care Unit, Hospital Sant Joan de Déu, Barcelona, Spain. ^134^Gestion Integral en Salud, Guatemala. ^135^Department of Internal Medicine, Hôpital Erasme, Université Libre de Bruxelles, Brussels, Belgium. ^136^Immunodeficiencies Unit, Research Institute Hospital, Madrid, Spain. ^137^Primary Immunodeficiencies Unit, Pediatrics, University Hospital 12 de Octubre, Madrid, Spain; School of Medicine Complutense University of Madrid, Madrid, Spain. ^138^Genomics Division, Instituto Tecnológico y de Energías Renovables (ITER), Santa Cruz de Tenerife, Spain. ^139^Assistance Publique Hôpitaux de Paris, Paris, France. ^140^Ankara City Hospital, Ankara, Turkey. ^141^Department of Intensive Care, Hospital Universitari de Bellvitge, IDIBELL, Barcelona, Spain. ^142^Immunodeficiency Outpatient Clinic, Institute for Medical Immunology, FOCIS Center of Excellence, Charité Universitätsmedizin Berlin, Germany. ^143^Surgical Intensive Care Unit, University Hospitals Leuven, Leuven, Belgium. ^144^CNAG-CRG, Barcelona Institute of Science and Technology, Barcelona, Spain. ^145^Department of Internal Medicine, National Reference Center for Rare Systemic Autoimmune Diseases, AP-HP, APHP-CUP, Hôpital Cochin, Paris, France. ^146^Department of Paediatric Immunology and Pulmonology, Center for Primary Immunodeficiency Ghent, Jeffrey Modell Diagnosis and Research Center, PID Research Lab, Ghent University Hospital, Ghent, Belgium. ^147^Sharjah Institute of Medical Research, College of Medicine, University of Sharjah, Sharjah, UAE, Prince Naif center for Immunology Research, King Saud University, Riyadh, SA. ^148^Division of Immunology, Department of Medical Biochemistry and Biophysics, Karolinska Institutet, Stockholm, Sweden. ^149^Department of Pediatrics (Infectious Diseases), Istanbul Faculty of Medicine, Istanbul University, Istanbul, Turkey. ^150^I. Horbachevsky Ternopil National Medical University, Ternopil, Ukraine. ^151^Pediatric Infectious Diseases Unit, Bakirkoy Dr. Sadi Konuk Training and Research Hospital, University of Health Sciences, Istanbul, Turkey. ^152^Health Sciences University, Darıca Farabi Education and Research Hospital, Kocaeli, Turkey. ^153^Department of Immunology, Hospital Universitario de Gran Canaria Dr. Negrín, Canarian Health System, Las Palmas de Gran Canaria, Spain. ^154^Department of Paediatrics, Queen Elizabeth Hospital, Hong Kong, China. ^155^Intensive Care Unit. Marqués de Valdecilla Hospital, Santander, Spain. ^156^Hospital del Mar, Institut Hospital del Mar d’Investigacions Mèdiques (IMIM), UAB, UPF, Barcelona, Spain. ^157^Intensive Care Unit, APHM, Marseille, France. ^158^CHU Lille, Lille, France. ^159^Department of Medicine, University of Hong Kong, Hong Kong, China. ^160^Department of Pediatrics, Columbia University, New York, NY, USA. ^161^Centre hospitalier intercommunal Poissy Saint Germain en Laye, Poissy, France. ^162^IHU Méditerranée Infection, Service de l’Information Médicale, Hôpital de la Timone, Marseille, France. ^163^Health Science University Ankara City Hospital, Ankara, Turkey. ^164^Eskişehir Osmangazi University, Pediatric Infectious Diseases, Eskişehir, Turkey. ^165^Mersin City Education and Research Hospital, Mersin, Turkey. ^166^Division of Pediatric Infectious Diseases, Prof. Dr. Cemil Tascıoglu City Hospital, Istanbul, Turkey. ^167^Departments of Infectious Diseases and Clinical Microbiology, Bakirkoy Dr. Sadi Konuk Training and Research Hospital, University of Health Sciences, Istanbul, Turkey. ^168^Department of Pediatric Rheumatology, Istanbul University-Cerrahpasa, Istanbul, Turkey. ^169^Department of Pediatrics, Tokyo Medical and Dental University, Tokyo, Japan. ^170^Health Sciences University, Umraniye Education and Research Hospital, Istanbul, Turkey. ^171^Department of Parasitology and Research Center for Infectious Disease Sciences, Graduate School of Medicine, Osaka City University, Osaka, Japan. ^172^Pediatric Infectious Diseases Unit of Osman Gazi University Medical School in Eskişehir, Turkey. ^173^Meram Medical Faculty, Necmettin Erbakan University, Konya, Turkey. ^174^Department of Immunology, Second Faculty of Medicine, Charles University and University Hospital in Motol, Prague, Czech Republic. ^175^ICU, First Department of Respiratory Medicine, National and Kapodistrian University of Athens, Medical School, “Sotiria” General Hospital of Chest Diseases, Athens, Greece. ^176^Central Clinical Hospital of the Ministry of Interior and Administration, Warsaw, Poland. ^177^Clinique des soins intensifs, HFR Fribourg, Fribourg, Switzerland. ^178^Oncobiologie Génétique Bioinformatique, PC Bio, CHU Besançon, Besançon, France. ^179^Department of Intensive Care, Tuen Mun Hospital, Hong Kong, China. ^180^Paediatric Infectious Disease Unit, Hospital Authority Infectious Disease Center, Princess Margaret Hospital, Hong Kong (Special Administrative Region), China. ^181^Department of Pathology, Queen Mary Hospital, Hong Kong, China. ^182^Aix Marseille Univ, IRD, MEPHI, IHU Méditerranée Infection, Marseille, France. ^183^Department of Paediatrics, Tuen Mun Hospital, Hong Kong, China. ^184^Biomedical Research Foundation of the Academy of Athens, Athens, Greece. ^185^Necker Hospital, Paris, France. ^186^Department of Paediatrics and Adolescent Medicine, University of Hong Kong, Hong Kong, China. ^187^National Centre for Infectious Diseases, Singapore, Singapore. ^188^Hospital Universitario Reina Sofía, Cordoba, Spain. ^189^Imperial College, London, England. ^190^Hospital General San Juan de Dios, Ciudad de Guatemala, Guatemala. ^191^Endocrinology and Diabetes for Children, AP-HP, Bicêtre Paris-saclay hospital, Le Kremlin-Bicêtre, France. ^192^Innate Immunity Group, IdiPAZ Institute for Health Research, La Paz Hospital, Madrid, Spain. ^193^Neurology Unit, AP-HP Pitié-Salpêtrière Hospital, Paris University, Paris, France. ^194^Department of Medicine, Pamela Youde Nethersole Eastern Hospital, Hong Kong, China. ^195^Intensive Care Unit, AP-HP Pitié-Salpêtrière Hospital, Paris University, Paris, France. ^196^National Centre for Infectious Diseases, Singapore, Singapore; Tan Tock Seng Hospital, Singapore, Singapore; Yong Loo Lin School of Medicine, Singapore, Singapore; Lee Kong Chian School of Medicine, Singapore, Singapore. ^197^Hospital de Niños Dr. Ricardo Gutierrez, Buenos Aires, Argentina. ^198^Department of Clinical Immunology and Infectious Diseases, National Research Institute of Tuberculosis and Lung Diseases, Shahid Beheshti University of Medical Sciences, Tehran, Iran. ^199^Neurooncology and Neuroinflammation Unit, IRCCS Mondino Foundation, Pavia, Italy. ^200^Clinical Tuberculosis and Epidemiology Research Center, National Research Institute of Tuberculosis and Lung Diseases (NRITLD), Shahid Beheshti University of Medical Sciences, Tehran, Iran. ^201^Coordenadora da Unidade de Infeciologia e Imunodeficiências do Serviço de Pediatria, Centro Materno-Infantil do Norte, Porto, Portugal. ^202^Hospital Sant Joan de Déu and University of Barcelona, Barcelona, Spain. ^203^Pediatric Infectious Diseases and Immunodeficiencies Unit, Hospital Universitari Vall d’Hebron, Vall d’Hebron Research Institute, Vall d’Hebron Barcelona Hospital Campus, Universitat Autònoma de Barcelona (UAB), Barcelona, Catalonia, Spain. ^204^Hospital Universitari Mutua de Terrassa, Universitat de Barcelona, Barcelona, Spain. ^205^IrsiCaixa AIDS Research Institute, ICREA, UVic-UCC, Research Institute “Germans Trias i Pujol,” Badalona, Spain. ^206^Department of Laboratory, Cruces University Hospital, Barakaldo, Bizkaia, Spain, Bizkaia, Spain. ^207^Intensive Care Unit, Hospital General Universitario “Gregorio Marañón,” Madrid, Spain. ^208^University of New South Wales, Sydney, NSW, Australia. ^209^AP-HP Pitié-Salpêtrière Hospital, Paris, France. ^210^Department of Pediatrics, Complejo Hospitalario Universitario Insular-Materno Infantil, Canarian Health System, Las Palmas de Gran Canaria, Spain. ^211^Medical Intensive Care Unit, University Hospitals Leuven, Leuven, Belgium. ^212^Aix-Marseille University, APHM, Marseille, France. ^213^Service de Médecine Intensive Réanimation, Hôpitaux Universitaires Henri Mondor, Assistance Publique–Hôpitaux de Paris (AP-HP), Groupe de Recherche Clinique CARMAS, Faculté de Santé de Créteil, Université Paris Est Créteil, France. ^214^AP-HP Cohin Hospital, Paris, France. ^215^Department of Critical Care Medicine, Ente Ospedaliero Cantonale, Bellinzona, Switzerland. ^216^Necmettin Erbakan University, Meram Medical Faculty, Division of Pediatric Infectious Diseases, Konya, Turke4y. ^217^Department of Pediatrics, University Hospitals Leuven, Leuven, Belgium; KU Leuven, Department of Microbiology, Immunology and Transplantation; Laboratory for Inborn Errors of Immunity, KU Leuven, Leuven, Belgium. ^218^Hospices Civils de Lyon, Hôpital de la Croix-Rousse, Lyon, France. ^219^Hôpital Erasme, Brussels, Belgium. ^220^Centre hospitalier de gonesse, Gonesse, France. ^221^Aix Marseille Univ, IRD, AP-HM, MEPHI, IHU Méditerranée Infection, Marseille, France. ^222^Vascular Medicine, Georges Pompidou Hospital, AP-HP, Paris, France. ^223^Institut Jérôme Lejeune, Paris, France. ^224^Division of Pulmonary and Critical Care, College of Medicine-Jacksonville, University of Florida, Jacksonville, FL, USA. ^225^Department of Pediatrics, Hiroshima University Graduate School of Biomedical and Health Sciences, Hiroshima, Japan. ^226^BC Children’s Hospital Research Institute, University of British Columbia, Vancouver, BC, Canada. ^227^Médecine Intensive Réanimation, Hôpitaux Universitaires Henri Mondor, Assistance Publique–Hôpitaux de Paris (AP-HP), Créteil, France. ^228^Guanarteme Health Care Center, Canarian Health System, Las Palmas de Gran Canaria, Spain. ^229^Regional University Hospital of Malaga, Malaga, Spain. ^230^Department of Immunology, Hospital Universitari de Bellvitge, IDIBELL, Barcelona, Spain. ^231^Aix Marseille Univ, INSERM, INRAE, C2VN, Marseille, France. ^232^Department of General Paediatrics, Hôpital Bicêtre, AP-HP, University of Paris Saclay, Le Kremlin-Bicêtre, France. ^233^INSERM U1144, Université de Paris, DMU INVICTUS, AP-HP.Nord, Département de Médecine Interne, Lariboisière Hospital, Paris, France. ^234^CHU de La Timone, Marseille, France. ^235^Department of Epidemiology, Infectious Disease Control and Prevention, Graduate School of Biomedical and Health Sciences, Hiroshima University, Hiroshima, Japan. ^236^Pediatric Immunology and Rheumatology Department, Necker Hospital, AP-HP, Paris, France. ^237^Centro Hospitalar Universitário de Lisboa Central, Lisbon, Portugal. ^238^Infectious Disease Horizontal Technology Centre, A*STAR, Singapore, Singapore; Singapore Immunology Network, A*STAR, Singapore. ^239^Department of Medicine and Geriatrics, Tuen Mun Hospital, Hong Kong, China. ^240^Regional Universitary Hospital of Malaga, Málaga, Spain. ^241^Department of Immunology, Hospital Universitario Marqués de Valdecilla, Santander, Spain. ^242^Bilkent University, Department of Molecular Biology and Genetics, Ankara, Turkey. ^243^BRFAA, Athens, Greece. ^244^IHU Méditerranée Infection, Aix Marseille Univ, IRD, AP-HM, SSA, VITROME, IHU Méditerranée Infection, Marseille, France. ^245^L’Hôpital Foch, Suresnes, France. ^246^Department of Immunology, CIC1408, GIMAP CIRI INSERM U1111, University Hospital of Saint-Etienne, Saint-Etienne, France. ^247^Department of Immunology, Hospital Universitario 12 de Octubre, Instituto de Investigación Sanitaria Hospital 12 de Octubre (imas12), Madrid, Spain. ^248^Unit of Biochemistry, Instituto Nacional de Ciencias Médicas y Nutrición Salvador Zubirán, Mexico City, Mexico. ^249^Diabetes Research Institute, IRCCS San Raffaele Hospital, Milan, Italy. ^250^AP-HP Hôpitaux Universitaires Paris-Sud, Le Kremlin-Bicêtre, France. ^251^AP-HP, Avicenne Hospital, Intensive Care Unit, Bobigny, France; INSERM UMR-S 942, Cardiovascular Markers in Stress Conditions (MASCOT), University of Paris, Paris, France. ^252^Neurometabolic Diseases Laboratory, IDIBELL-Hospital Duran i Reynals, Barcelona; CIBERER U759, ISCiii Madrid, Spain. ^253^Hospices Civils de Lyon, Lyon, France. ^254^Univ. Lille, INSERM U1285, CHU Lille, Pôle de médecine intensive-réanimation, CNRS, UMR 8576–Unité de Glycobiologie Structurale et Fonctionnelle, Lille, France. ^255^Department of General pediatrics, Robert Debre Hospital, Paris, France. ^256^University of British Columbia, Vancouver, BC, Canada. ^257^Jeffrey Modell Diagnostic and Research Center for Primary Immunodeficiencies, Barcelona, Catalonia, Spain; Diagnostic Immunology Research Group, Vall d’Hebron Research Institute (VHIR), Vall d’Hebron University Hospital (HUVH), Vall d’Hebron Barcelona Hospital Campus, Barcelona, Catalonia, Spain. ^258^AP-HP, Avicenne Hospital, Intensive Care Unit, Bobigny, France; University Sorbonne Paris Nord, Bobigny, France. ^259^Centre Hospitalier de Saint-Denis, St Denis, France. ^260^Precision Medicine Unit, Lausanne University Hospital and University of Lausanne, Lausanne, Switzerland. ^261^Paris Cardiovascular Center, PARCC, INSERM, Université de Paris, Paris, France. ^262^Germans Trias i Pujol Hospital, Badalona, Spain. ^263^Medical Intensive Care Unit, Hopital de la Croix-Rousse, Hospices Civils de Lyon, Lyon, France. ^264^Department of Clinical Laboratory, Hospital Universitari de Bellvitge, IDIBELL, Barcelona, Spain. ^265^Pediatric Infectious Diseases and Immunodeficiencies Unit, Hospital Universitari Vall d’Hebron, Vall d’Hebron Research Institute, Vall d’Hebron Barcelona Hospital Campus., Barcelona, Spain. ^266^Université de Paris, CNRS UMR-8601, Paris, France; Team Chemistry and Biology, Modeling and Immunology for Therapy, CBMIT, Paris, France. ^267^Germans Trias i Pujol University Hospital and Research Institute, Badalona, Spain. ^268^Department of Immunology, University Hospital of Gran Canaria Dr. Negrín, Canarian Health System, Las Palmas de Gran Canaria, Spain; Department of Clinical Sciences, University Fernando Pessoa Canarias, Las Palmas de Gran Canaria, Spain. ^269^Neurometabolic Diseases Laboratory, Bellvitge Biomedical Research Institute (IDIBELL), 08908 L’Hospitalet de Llobregat, Barcelona, Spain; University Hospital Germans Trias i Pujol, Badalona, Barcelona, Catalonia, Spain. ^270^Consorcio Hospital General Universitario, Valencia, Spain. ^271^AP-HP Hôpitaux Universitaires Paris-Sud, Paris, France. ^272^Intensive Care Unit, Louis-Mourier Hospital, Colombes, France. ^273^Virology Unit, Université de Paris, Cohin Hospital, AP-HP, Paris, France. ^274^Neurometabolic Diseases Laboratory and CIBERER U759, Barcelona, Spain. ^275^Hospital San Pedro, Logroño, Spain. ^276^University of Tartu, Institute of Biomedicine and Translational Medicine, Tartu, Estonia. ^277^Respiratory Medicine, Georges Pompidou Hospital, AP-HP, Paris, France. ^278^Infectious Diseases Department, International Health Program of the Catalan Institute of Health (PROSICS), Vall d’Hebron University Hospital (HUVH), Vall d’Hebron Barcelona Hospital Campus, Universitat Autónoma de Barcelona, Barcelona, Spain. ^279^Hospital Clínico San Carlos and IdSSC, Madrid, Spain. ^280^Faculdades Pequeno Príncipe, Instituto de Pesquisa Pelé Pequeno Príncipe, Curitiba, Brazil. ^281^AP-HP, Avicenne Hospital, Intensive Care Unit, Bobigny, France. ^282^Service de Médecine Intensive Réanimation, Institut de Cardiologie, Hopital Pitié-Salpêtrière, Paris, France. ^283^Department of Pediatrics, Medizinische Fakultät Carl Gustav Carus, Technische Universität Dresden, Dresden, Germany. ^284^CHRU de Nancy, Hôpital d’Enfants, Vandoeuvre, France. ^285^Chair of Nephrology, University of Brescia, Brescia, Italy. ^286^Department of Immunology, Second Faculty of Medicine, Charles University and Motol University Hospital, Prague, Czech Republic. ^287^Clínica Universidad de Navarra and Ciberes, Madrid, Spain. ^288^HUS Helsinki University Hospital, Children and Adolescents, Rare Disease Center, and Inflammation Center, Adult Immunodeficiency Unit, Majakka, Helsinki, Finland. ^289^Fundació Docencia i Recerca Mutua Terrassa, Terrassa, Spain. ^290^D. Rogachev National Medical and Research Center of Pediatric Hematology, Oncology, Immunoogy, Moscow, Russia. ^291^Haematology Laboratory, Lariboisière Hospital, University of Paris, Paris, France. ^292^Biomedical Research Foundation of the Academy of Athens, Athens, Greece. ^293^INSERM U1140, University of Paris, European Georges Pompidou Hospital, Paris, France. ^294^Department of Pediatrics, Faculty of Medicine, Mansoura University, Mansoura, Egypt. ^295^Intensive Care Medicine, Hospital Universitario de Gran Canaria Dr. Negrín, Canarian Health System, Las Palmas de Gran Canaria, Spain. ^296^CHU de Saint Etienne, Saint-Priest-en-Jarez, France. ^297^Pediatric Infectious Diseases and Immunodeficiencies Unit, Hospital Universitari Vall d’Hebron, Vall d’Hebron Research Institute, Vall d’Hebron Barcelona Hospital Campus. Universitat Autònoma de Barcelona (UAB), Barcelona, Catalonia, Spain. ^298^Department of Pediatric Infectious Diseases and Pediatric Immunology, Shupyk National Healthcare University of Ukraine, Kyiv, Ukraine. ^299^Gustave Roussy Cancer Campus, Villejuif, France. ^300^Intensive Care Unit, Avicenne Hospital, AP-HP, Bobigny, France. ^301^Laboratory of Immunology and Histocompatibility, Saint-Louis Hospital, Paris University, Paris, France. ^302^Center for Inflammation Research, Laboratory of Molecular Signal Transduction in Inflammation, VIB, Ghent, Belgium. ^303^Department of Internal Medicine, Université de Paris, INSERM, U970, PARCC, F-75015, Paris, France. ^304^Service de médecine intensive réanimation, CHU de Saint-Etienne, France. ^305^Department of Rheumatology and Inflammation Research, Institute of Medicine, Sahlgrenska Academy, University of Gothenburg, Gothenburg, Sweden. ^306^University of Management and Technology, Lahore, Pakistan. ^307^Department of Infectious Diseases, Aarhus University Hospital, Aarhus, Denmark. ^308^First Division of Anesthesiology and Critical Care Medicine, University of Brescia, ASST Spedali Civili di Brescia, Brescia, Italy. ^309^Intensive Care Department, Hospital Universitari MutuaTerrassa, Universitat Barcelona, Terrassa, Spain. ^310^Laboratory of Immunobiology, Center for Clinical, Experimental Surgery and Translational Research, Biomedical Research Foundation of the Academy of Athens, Athens, Greece. ^311^International Center of Research in Infectiology, Lyon University, INSERM U1111, CNRS UMR 5308, ENS, UCBL, Lyon, France; Hospices Civils de Lyon, Lyon Sud Hospital, Pierre-Bénite, France. ^312^Infanta Leonor University Hospital, Madrid, Spain. ^313^Department of Medicine and Geriatrics, Princess Margaret Hospital, Hong Kong, China. ^314^University of Tartu, Institute of Clinical Medicine, Tartu, Estonia. ^315^Department of Medicine, United Christian Hospital, Hong Kong, China. ^316^Hematology Department, ASST Spedali Civili di Brescia, Brescia, Italy. ^317^Pneumologie, Hôpital Avicenne, AP-HP, INSERM U1272, Université Sorbonne Paris Nord, Bobigny, France. ^318^Dermatology Unit, Laboratoire GAD, INSERM UMR1231 LNC, Université de Bourgogne, Dijon, France. ^319^University Hospital of Burgos, Burgos, Spain. ^320^Center of Human Genetics, Hôpital Erasme, Université Libre de Bruxelles, Brussels, Belgium. ^321^Bellvitge University Hospital, L’Hospitalet de Llobregat, Barcelona, Spain. ^322^University of São Paulo, São Paulo, Brazil. ^323^CHU de Caen, Caen, France. ^324^Hospital del Mar–IMIM Biomedical Research Institute, Barcelona, Catalonia, Spain. ^325^Pediatric Infectious Diseases, Pediatric Department, Cliniques universitaires Saint-Luc, UCL Louvain, Brussels Belgium. ^326^Sorbonne Université, Service de Médecine Intensive Réanimation, Hôpital Tenon, Assistance Publique-Hôpitaux de Paris, Paris, France. ^327^Pediatric Infectious Disease and Pediatric Immunology Department, Shupyk National Healthcare University of Ukraine, Kyiv, Ukraine. ^328^Department of Pneumology, University Hospitals Leuven, Leuven, Belgium. ^329^Laboratory for Clinical Infectious and Inflammatory Disorders, Department of Microbiology, Immunology, and Transplantation, Leuven, Belgium. ^330^Department of Clinical Pathology, Pamela Youde Nethersole Eastern Hospital, Hong Kong, China. ^331^Department of Medicine, Queen Elizabeth Hospital, Hong Kong, China. ^332^Ankara City Hospital, Children’s Hospital, Ankara, Turkey. ^333^Division of Pediatric Infectious Disease, Department of Pediatrics, Faculty of Medicine, Karadeniz Technical University, Trabzon, Turkey. ^334^Health Sciences University, Lütfi Kırdar Kartal Education and Research Hospital, İstanbul, Turkey. ^335^Department of Nephrology and Infectiology, AZ Sint-Jan, Bruges, Belgium. ^336^Department of Respiratory Medicine, Ghent University Hospital, Belgium. ^337^Department of Pediatric Pulmonology and Immunology, Ghent University Hospital, Ghent, Belgium. ^338^Department of Intensive Care Unit, Ghent University Hospital, Ghent, Belgium. ^339^Department of Pediatric Hemato-oncology, Jolimont Hospital, La louvière, Belgium. ^340^Department of Pulmonology, ZNA Middelheim, Antwerp, Belgium. ^341^Department of Internal Medicine, Ghent University Hospital, Ghent, Belgium. ^342^Department of Pediatric Immuno-hémato-rheumatology, CHR Citadelle, Liége, Belgium. ^343^Intensive Care Unit, Grand Hôpital de l’Est Francilien Site de Marne-La-Vallée, Jossigny, France. ^344^Clinical Translational and Engagement Platform, Garvan Institute of Medical Research, Darlinghurst, NSW, Australia; School of Clinical Medicine, UNSW Medicine & Health, St Vincent’s Healthcare Clinical Campus, Faculty of Medicine and Health, UNSW Sydney, Kensington, NSW, Australia. ^345^Department of Immunology, University Hospital Ramón y Cajal, Madrid, Spain. ^346^Respiratory Diseases Division, IRCCS Policlinico San Matteo Foundation, University of Pavia, Pavia, Italy.

***Members of Orchestra Working Group*:** Laurent Abel^1^, Matilda Berkell^2^, Valerio Carelli^3^, Alessia Fiorentino^3^, Surbhi Malhotra^2^, Alessandro Mattiaccio^3^, Tommaso Pippucci^3^, Marco Seri^3^, Evelina Tacconelli^4^

^1^Inserm, University Paris cité, Imagine Institute, Paris, France, ^2^University of Antwerp, Antwerp, Belgium, ^3^University of Bologna, Bologna, 40138, Italy, ^4^University of Verona, 37129 Verona, Italy

***Members of French COVID Cohort Study Group*:** Laurent Abel^1^, Claire Andrejak^2^, François Angoulvant^3^, Delphine Bachelet^4^, Marie Bartoli^5^, Romain Basmaci^6^, Sylvie Behillil^7^, Marine Beluze^8^, Dehbia Benkerrou^9^, Krishna Bhavsar^4^, Lila Bouadma^4^, Sabelline Bouchez^10^, Maude Bouscambert^11^, Minerva Cervantes-Gonzalez^4^, Anissa Chair^4^, Catherine Chirouze^12^, Alexandra Coelho^13^, Camille Couffignal^4^, Sandrine Couffin-Cadiergues^14^, Eric d’Ortenzio^5^, Marie-Pierre Debray^4^, Laurene Deconinck^4^, Dominique Deplanque^15^, Diane Descamps^4^, Mathilde Desvallée^16^, Alpha Diallo^5^, Alphonsine Diouf^13^, Céline Dorival^9^, François Dubos^17^, Xavier Duval^4^, Brigitte Elharrar^18^, Philippine Eloy^4^, Vincent Enouf^7^, Hélène Esperou^14^, Marina Esposito-Farese^4^, Manuel Etienne^19^, Eglantine Ferrand Devouge^19^, Nathalie Gault^4^, Alexandre Gaymard^11^, Jade Ghosn^4^, Tristan Gigante^20^, Morgane Gilg^20^, Jérémie Guedj^21^, Alexandre Hoctin^13^, Isabelle Hoffmann^4^, Ikram Houas^14^, Jean-Sébastien Hulot^22^, Salma Jaafoura^14^, Ouifiya Kafif^4^, Florentia Kaguelidou^23^, Sabrina Kali^4^, Antoine Khalil^4^, Coralie Khan^16^, Cédric Laouénan^4^, Samira Laribi^4^, Minh Le^4^, Quentin Le Hingrat^4^, Soizic Le Mestre^5^, Hervé Le Nagard^24^, François-Xavier Lescure^4^, Sophie Letrou^4^, Yves Levy^25^, Bruno Lina^11^, Guillaume Lingas^24^, Jean Christophe Lucet^4^, Denis Malvy^26^, Marina Mambert^13^, France Mentré^4^, Amina Meziane^9^, Hugo Mouquet^7^, Jimmy Mullaert^4^, Nadège Neant^24^, Duc Nguyen^26^, Marion Noret^27^, Saad Nseir^17^, Aurélie Papadopoulos^14^, Christelle Paul^5^, Nathan Peiffer-Smadja^4^, Thomas Perpoint^28^, Ventzislava Petrov-Sanchez^5^, Gilles Peytavin^4^, Huong Pham^4^, Olivier Picone^6^, Valentine Piquard^4^, Oriane Puéchal^29^, Christian Rabaud^30^, Manuel Rosa-Calatrava^11^, Bénédicte Rossignol^20^, Patrick Rossignol^30^, Carine Roy^4^, Marion Schneider^4^, Richa Su^4^, Coralie Tardivon^4^, Marie-Capucine Tellier^4^, François Téoulé^9^, Olivier Terrier^11^, Jean-François Timsit^4^, Christelle Tual^31^, Sarah Tubiana^4^, Sylvie Van Der Werf^7^, Noémie Vanel^32^, Aurélie Veislinger^31^, Benoit Visseaux^4^, Aurélie Wiedemann^25^, Yazdan Yazdanpanah^4^

^1^INSERM UMR 1163, Paris, France. ^2^CHU Amiens, Amiens, France. ^3^Hôpital Necker, Paris, France. ^4^Hôpital Bichat, Paris, France. ^5^ANRS, Paris, France. ^6^Hôpital Louis Mourier, Colombes, France. ^7^Pasteur Institute, Paris, France. ^8^F-CRIN Partners Platform, Paris, France. ^9^INSERM UMR 1136, Paris, France. ^10^CHU Nantes, France. ^11^INSERM UMR 1111, Lyon, France. ^12^CHRU Jean Minjoz, Besançon, France. ^13^INSERM UMR 1018, Paris, France. ^14^INSERM Sponsor, Paris, France. ^15^Centre d’Investigation Clinique, INSERM CIC 1403, Centre Hospitalo universitaire de Lille, Lille, France. ^16^INSERM UMR 1219, Bordeaux, France. ^17^CHU Lille, Lille, France. ^18^CHI de Créteil, Créteil, France. ^19^CHU Rouen, Rouen, France. ^20^F-CRIN INI-CRCT, Nancy, France. ^21^Université de Paris, INSERM, IAME, F-75018 Paris, France. ^22^Hôpital Européen Georges Pompidou, Paris, France. ^23^Hôpital Robert Debré, Paris, France. ^24^INSERM UMR 1137, Paris, France. ^25^Vaccine Research Institute (VRI), INSERM UMR 955, Créteil, France. ^26^CHU Bordeaux, Bordeaux, France. ^27^RENARCI, Annecy, France. ^28^CHU Lyon, Lyon, France. ^29^REACTing, Paris, France. ^30^CHU Nancy, Nancy, France. ^31^INSERM CIC-1414, Rennes, France. ^32^Hôpital la Timone, Marseille, France.

***Members of CoV-Contact Cohort*:** Loubna Alavoine^1^, Sylvie Behillil^2^, Charles Burdet^3^, Charlotte Charpentier^4^, Aline Dechanet^5^, Diane Descamps^6^, Xavier Duval^7^, Jean-Luc Ecobichon^1^, Vincent Enouf^8^, Wahiba Frezouls^1^, Nadhira Houhou^5^, Ouifiya Kafif^5^, Jonathan Lehacaut^1^, Sophie Letrou^1^, Bruno Lina^9^, Jean-Christophe Lucet^10^, Pauline Manchon^5^, Mariama Nouroudine^1^, Valentine Piquard^5^, Caroline Quintin^1^, Michael Thy^11^, Sarah Tubiana^1^, Sylvie van der Werf^8^, Valérie Vignali^1^, Benoit Visseaux^10^, Yazdan Yazdanpanah^10^, Abir Chahine^12^, Nawal Waucquier^12^, Maria-Claire Migaud^12^, Dominique Deplanque^12^, Félix Djossou^13^, Mayka Mergeay-Fabre^14^, Aude Lucarelli^15^, Magalie Demar^13^, Léa Bruneau^16^, Patrick Gérardin^17^, Adrien Maillot^16^, Christine Payet^18^, Bruno Laviolle^19^, Fabrice Laine^19^, Christophe Paris^19^, Mireille Desille-Dugast^19^, Julie Fouchard^19^, Denis Malvy^20^, Duc Nguyen^20^, Thierry Pistone^20^, Pauline Perreau^20^, Valérie Gissot^21^, Carole L. E. Goas^21^, Samatha Montagne^22^, Lucie Richard^23^, Catherine Chirouze^24^, Kévin Bouiller^24^, Maxime Desmarets^25^, Alexandre Meunier^26^, Marilou Bourgeon^26^, Benjamin Lefévre^27^, Hélène Jeulin^28^, Karine Legrand^29^, Sandra Lomazzi^30^, Bernard Tardy^31^, Amandine Gagneux-Brunon^32^, Frédérique Bertholon^33^, Elisabeth Botelho-Nevers^32^, Christelle Kouakam^34^, Leturque Nicolas^34^, Layidé Roufai^34^, Karine Amat^35^, Sandrine Couffin-Cadiergues^34^, Hélène Espérou^36^, Samia Hendou^34^

^1^Centre d’Investigation Clinique, INSERM CIC 1425, Hôpital Bichat Claude Bernard, AP-HP, Paris, France. ^2^Institut Pasteur, Paris, France. ^3^Université de Paris, IAME, INSERM U1137, Paris, France; Hôpital Bichat Claude Bernard, AP-HP, Paris, France. ^4^Service de Virologie, Université de Paris, INSERM, IAME, UMR 1137, Hôpital Bichat Claude Bernard, AP-HP, Paris, France. ^5^Hôpital Bichat Claude Bernard, AP-HP, Paris, France. ^6^IAME INSERM U1140, Hôpital Bichat Claude Bernard, AP-HP, Paris, France. ^7^Centre d’Investigation Clinique, INSERM CIC 1425, AP-HP, IAME, Paris University, Paris, France. ^8^Institut Pasteur, U3569 CNRS, Université de Paris, Paris, France. ^9^Virpath Laboratory, International Center of Research in Infectiology, Lyon University, INSERM U1111, CNRS U5308, ENS, UCBL, Lyon, France. ^10^IAME INSERM U1138, Hôpital Bichat Claude Bernard, AP-HP, Paris, France. ^11^Center for Clinical Investigation, Assistance Publique-Hôpitaux de Paris, Bichat-Claude Bernard University Hospital, Paris, France. ^12^Centre d’Investigation Clinique, INSERM CIC 1403, Centre Hospitalo universitaire de Lille, Lille, France. ^13^Service des maladies infectieuses, Centre Hospitalo universitaire de Cayenne, Guyane, France. ^14^Centre d’Investigation Clinique, INSERM CIC 1424, Centre Hospitalier de Cayenne, Cayenne, Guyane Française. ^15^Service Hôpital de jour Adulte, Centre Hospitalier de Cayenne, Guyane, France. ^16^Centre d’Investigation Clinique, INSERM CIC 1410, Centre Hospitalo universitaire de la Réunion, La Réunion, France. ^17^Centre d’Investigation Clinique, INSERM CIC 1410, CHU Reunion, Saint-Pierre, Reunion Island. ^18^Centre d’Investigation Clinique, INSERM CIC 1410, Centre de Ressources Biologiques, Centre Hospitalo universitaire de la Réunion, La Réunion, France. ^19^Centre d’Investigation Clinique, INSERM CIC 1414, Centre Hospitalo universitaire de Rennes, Rennes, France; Centre de Ressources Biologiques, CRB Santé, Centre Hospitalo universitaire de Rennes, Rennes, France. ^20^Service des maladies infectieuses, Centre Hospitalo universitaire de Bordeaux, Bordeaux, France. ^21^Centre d’Investigation Clinique, INSERM CIC 1415, CHRU Tours, Tours, France. ^22^CRBT, Centre Hospitalo universitaire de Tours, Tours, France. ^23^Pole de Biologie Médicale, Centre Hospitalo universitaire de Tours, Tours, France. ^24^Service des maladies infectieuses, Centre Hospitalo universitaire de Besançon, Besançon, France. ^25^Service des maladies infectieuses, Centre d’investigation clinique, INSERM CIC1431, Centre Hospitalier Universitaire de Besançon, Besançon, France. ^26^Centre de Ressources Biologiques–Filière Microbiologique de Besançon, Centre Hospitalier Universitaire, Besançon, France. ^27^Université de Lorraine, CHRU-Nancy and APEMAC, Infectious and Tropical Diseases, Nancy, France. ^28^Laboratoire de Virologie, CHRU de Nancy Brabois, Vandoeuvre-lès-Nancy, France. ^29^INSERM CIC-EC 1433, Centre Hospitalo universitaire de Nancy, Nancy, France. ^30^Centre de ressources Biologiques, Centre Hospitalo universitaire de Nancy, Nancy, France. ^31^Centre d’Investigation Clinique, INSERM CIC 1408, Centre Hospitalo universitaire de Saint Etienne, Saint Etienne, France. ^32^Service des maladies infectieuses, Centre Hospitalo universitaire de Saint Etienne, Saint Etienne, France. ^33^Service des maladies infectieuses, CRB^42^-BTK, Centre Hospitalo Universitaire de Saint Etienne, Saint Etienne, France. ^34^Pole Recherche Clinique, INSERM, Paris, France. ^35^IMEA Fondation Léon M’Ba, Paris, France. ^36^INSERM Clinical Research Department, Paris, France.

***Members of the COVIDeF study group*:** Serge Bureau^1^, Yannick Vacher^1^, Anne Gysembergh-Houal^1^, Lauren Demerville^1^, Abla Chachoua^1^, Sebastien Abad^2^, Radhiya Abassi^3^, Abdelrafie Abdellaoui^3^, Abdelkrim Abdelmalek^4^, Hendy Abdoul^5^, Helene Abergel^6^, Fariza Abeud^7^, Sophie Abgrall^8^, Noemie Abisror^4^, Marylise Adechian^9^, Nordine Aderdour^9^, Hakeem Farid Admane^4^, Frederic Adnet^2^, Sara Afritt^5^, Helene Agostini^10^, Claire Aguilar^11^, Sophie Agut^12^, Tommaso Francesco Aiello^13^, Marc Ait Kaci^14^, Hafid Ait Oufella^4^, Gokula Ajeenthiravasan^15^, Virginie Alauzy^3^, Fanny Alby-Laurent^11^, Lucie Allard^2^, Marie-Alexandra Alyanakian^11^, Blanca Amador Borrero^7^, Sabrina Amam^6^, Lucile Amrouche^11^, Marc Andronikof^16^, Dany Anglicheau^11^, Nadia Anguel^9^, Djillali Annane^15^, Mohammed Aounzou^3^, Caroline Aparicio^7^, Gladys Aratus^4^, Jean-Benoit Arlet^14^, Jeremy Arzoine^3^, Elisabeth Aslangul^13^, Mona Assefi^3^, Adeline Aubry^3^, Laetitia Audiffred^4^, Etienne Audureau^17^, Christelle Nathalie Auger^5^, Jean-Charles Auregan^8^, Celine Awotar^11^, Sonia Ayllon Milla^5^, Delphine Azan^5^, Laurene Azemar^7^, Billal Azzouguen^7^, Marwa Bachir Elrufaai^12^, Aïda Badsi^7^, Prissile Bakouboula^11^, Coline Balcerowiak^3^, Fanta Balde^12^, Elodie Baldivia^17^, Eliane-Flore Bangamingo^18^, Amandine Baptiste^3^, Fanny Baran-Marszak^2^, Caroline Barau^17^, Nathalie Barget^19^, Flore Baronnet^3^, Romain Barthelemy^7^, Jean-Luc Baudel^4^, Camille Baudry^2^, Elodie Baudry^9^, Laurent Beaugerie^4^, Adel Belamri^3^, Nicolas Belaube^12^, Rhida Belilita^3^, Pierre Bellassen^3^, Rawan Belmokhtar^2^, Isabel Beltran^6^, Ruben Benainous^2^, Mourad Benallaoua^2^, Robert Benamouzig^2^, Amélie Benbara^19^, Jaouad Benhida^3^, Anis Benkhelouf^3^, Jihene Benlagha^18^, Chahinez Benmostafa^14^, Skander Benothmane^18^, Miassa Bentifraouine^2^, Laurence Berard^4^, Quentin Bernier^3^, Enora Berti^17^, Astrid Bertier^9^, Laure Berton^7^, Simon Bessis^15^, Alexandra Beurton^20^, Celine Bianco^4^, Clara Bianquis^3^, Frank Bidar^3^, Philippe Blanche^5^, Clarisse Blayau^12^, Alexandre Bleibtreu^3^, Emmanuelle Blin^12^, Coralie Bloch-Queyrat^2^, Marie-Christophe Boissier^2^, Diane Bollens^4^, Marion Bolzoni^4^, Rudy pierre Bompard^12^, Nicolas Bonnet^2^, Justine Bonnouvrier^4^, Shirmonecrystal Botha^3^, Wissam Boucenna^4^, Fatiha Bouchama^3^, Olivier Bouchaud^2^, Hanane Bouchghoul^9^, Taoueslylia Boudjebla^12^, Noel Boudjema^17^, Catherine Bouffard^6^, Adrien Bougle^3^, Meriem Bouguerra^3^, Leila Bouras^7^, Agnes Bourcier^3^, Anne Bourgarit Durand^19^, Anne Bourrier^4^, Fabrice Bouscarat^6^, Diane Bouvry^2^, Nesrine Bouziri^3^, Ons Bouzrara^3^, Sarah Bribier^9^, Delphine Brugier^3^, Melanie Brunel^11^, Eida Bui^4^, Anne Buisson^21^, Iryna Bukreyeva^9^, Côme Bureau^20^, Jacques Cadranel^12^, Johann Cailhol^2^, Ruxandra Calin^12^, Clara Campos Vega^11^, Pauline Canavaggio^3^, Marta Cancella^3^, Delphine Cantin^22^, Albert Cao^3^, Lionel Carbillon^19^, Nicolas Carlier^5^, Clementine Cassard^3^, Guylaine Castor^7^, Marion Cauchy^7^, Olivier Cha^4^, Benjamin Chaigne^5^, Salima Challal^2^, Karine Champion^7^, Patrick Chariot^19^, Julie Chas^12^, Simon Chauveau^2^, Anthony Chauvin^7^, Clement Chauvin^18^, Nathalie Chavarot^11^, Kamélia Chebbout^3^, Mustapha Cherai^3^, Ilaria Cherubini^3^, Amelie Chevalier^5^, Thibault Chiarabini^4^, Thierry Chinet^10^, Richard Chocron^14^, Pascaline Choinier^12^, Juliette Chommeloux^3^, Christophe Choquet^6^, Laure Choupeaux^11^, Benjamin Chousterman^7^, Dragosmarius Ciocan^8^, Ada Clarke^5^, Gaëlle Clavere^23^, Florian Clavier^3^, Karine Clement^3^, Sebastien Clerc^14^, Yves Cohen^2^, Fleur Cohen^3^, Adrien Cohen^23^, Audrey Coilly^24^, Hester Colboc^25^, Pauline Colin^3^, Magalie Collet^7^, Chloé Comarmond^7^, Emeline Combacon^5^, Alain Combes^3^, Celine Comparon^2^, Jean-Michel Constantin^3^, Hugues Cordel^2^, Anne-Gael Cordier^9^, Adrien Costantini^10^, Nathalie Costedoat Chalumeau^5^, Camille Couffignal^6^, Doriane Coupeau^4^, Alain Creange^17^, Yannie Cuvillier Lamarre^22^, Charlène Da Silveira^6^, Sandrine Dautheville Guibal El Kayani^12^, Nathalie De Castro^18^, Yann De Rycke^3^, Lucie Del Pozo^19^, Quentin Delannoy^3^, Mathieu Delay^12^, Robin Deleris^3^, Juliette Delforge^13^, Laëtitia Delphine^3^, Noemie Demare^2^, Sophie Demeret^3^, Alexandre Demoule^3^, Aurore Deniau^2^, François Depret^18^, Sophie Derolez^2^, Ouda Derradji^9^, Nawal Derridj^10^, Vincent Descamps^6^, Lydia Deschamps^6^, Celine Desconclois^8^, Cyrielle Desnos^3^, Karine Desongins^18^, Robin Dhote^2^, Benjamin Diallo^12^, Morgane Didier^2^, Myriam Diemer^7^, Stephane Diez^9^, Juliette Djadi-Prat^14^, Fatima-Zohra Djamouri Monnory^12^, Siham Djebara^3^, Naoual Djebra^2^, Minette Djietcheu^18^, Hadjer Djillali^4^, Nouara Djouadi^4^, Severine Donneger^19^, Catarina Dos Santos^5^, Nathalie Dournon^2^, Martin Dres^20^, Laura Droctove^3^, Marie Drogrey^3^, Margot Dropy^3^, Elodie Drouet^4^, Valérie Dubosq^12^, Evelyne Dubreucq^12^, Estelle Dubus^7^, Boris Duchemann^2^, Thibault Duchenoy^5^, Emmanuel Dudoignon^18^, Romain Dufau^19^, Florence Dumas^5^, Clara Duran^10^, Emmanuelle Duron^24^, Antoine Durrbach^17^, Claudine Duvivier^11^, Nathan Ebstein^2^, Jihane El Khalifa^6^, Alexandre Elabbadi^12^, Caroline Elie^11^, Gabriel Ernotte^3^, Anne Esling^11^, Martin Etienne^9^, Xavier Eyer^7^, Muriel sarah Fartoukh^12^, Takoua Fayali^3^, Marion Fermaut^19^, Arianna Fiorentino^4^, Souha Fliss^2^, Marie-Céline Fournier^7^, Benjamin Fournier^11^, Hélène Francois^12^, Olivia Freynet^2^, Yvann Frigout^14^, Isaure Fromont^7^, Axelle Fuentes^6^, Thomas Furet^3^, Joris Galand^7^, Marc Garnier^4^, Agnes Gaubert^3^, Stéphane Gaudry^2^, Samuel Gaugain^7^, Damien Gauthier^3^, Maxime Gautier^7^, Sophie Georgin-Lavialle^12^, Daniela Geromin^14^, Mohamed Ghalayini^2^, Bijan Ghaleh^17^, Myriam Ghezal^21^, Aude Gibelin^12^, Linda Gimeno^3^, Benoit Girard^5^, Bénédicte Giroux Leprieur^2^, Doryan Gomes^18^, Elisabete Gomes-Pires^11^, Guy Gorochov^3^, Anne Gouge^18^, Amel Gouja^17^, Helene Goulet^12^, Sylvain Goupil^11^, Jeanne Goupil De Bouille^2^, Julien Gras^7^, Segolene Greffe^10^, Lamiae Grimaldi^9^, Paul Guedeney^3^, Bertrand Guidet^4^, Matthias Guillo^18^, Mariechristelle Gulczynski^26^, Tassadit Hadjam^7^, Didier Haguenauer^13^, Soumeya Hammal^3^, Nadjib Hammoudi^3^, Olivier Hanon^23^, Anarole Harrois^9^, Pierre Hausfater^3^, Coraline Hautem^14^, Guillaume Hekimian^3^, Nicholas Heming^15^, Olivier Hermine^11^, Sylvie Ho^3^, Marie Houllier^9^, Benjamin Huot^7^, Tessa Huscenot^7^, Wafa Ibn Saied^12^, Ghilas Ikherbane^3^, Meriem Imarazene^11^, Patrick Ingiliz^4^, Lina Iratni^17^, Stephane Jaureguiberry^9^, Jean-Francois Jean-Marc^10^, Deleena Jeyarajasingham^18^, Pauline Jouany^14^, Veronique Jouis^7^, Clement Jourdaine^7^, Ouifiya Kafif^6^, Rim Kallala^24^, Sandrine Katsahian^14^, Lilit Kelesyan^27^, Vixra Keo^3^, Flora Ketz^21^, Warda Khamis^2^, Enfel Khelili^3^, Mehdi Khellaf^17^, Christy Gaëlla Kotokpo Youkou^10^, Ilias Kounis^24^, Gaelle Kpalma^3^, Jessica Krause^4^, Vincent Labbe^12^, Karine Lacombe^4^, Jean-Marc Lacorte^3^, Anne Gaelle Lafont^4^, Emmanuel Lafont^11^, Lynda Lagha^27^, Lionel Lamhaut^11^, Aymeric Lancelot^3^, Cecilia Landman^4^, Fanny Lanternier^11^, Cecile Larcheveque^3^, Caroline Lascoux Combe^18^, Ludovic Lassel^12^, Benjamin Laverdant^12^, Christophe Lavergne^18^, Jean-Rémi Lavillegrand^4^, Pompilia Lazureanu^7^, Loïc Le Guennec^3^, Lamia Leberre^4^, Claire Leblanc^19^, Marion Leboyer^28^, Francois Lecomte^5^, Marine Lecorre^3^, Romain Leenhardt^4^, Marylou Lefebvre^4^, Bénédicte Lefebvre^4^, Paul Legendre^5^, Anne Leger^3^, Laurence Legros^24^, Justyna Legrosse^3^, Sébastien Lehuunghia^5^, Julien Lemarec^3^, Jeremie Leporrier-Ext^11^, Manon Lesein^5^, Hubert Lesur^24^, Vincent Levy^2^, Albert Levy^14^, Edwige Lopes^7^, Amanda Lopes^7^, Vanessa Lopez^11^, Julien Lopinto^12^, Olivier Lortholary^11^, Badr Louadah^7^, Bénédicte Loze^18^, Marie-Laure Lucas^22^, Axelle Lucasamichi^8^, Liem Binh Luong^5^, Arouna Magazimama-Ext^7^, David Maingret^7^, Lakhdar Mameri^18^, Philippe Manivet^7^, Cylia Mansouri^4^, Estelle Marcault^6^, Jonathan Marey^5^, Nathalie Marin^5^, Clémence Marois^3^, Olivier Martin^2^, Lou Martineau^3^, Cannelle Martinez-Lopez^15^, Pierre Martyniuck^4^, Pauline Mary De Farcy^29^, Nessrine Marzouk^12^, Rafik Masmoudi^14^, Alexandre Mebazaa^7^, Frédéric Mechai^2^, Fabio Mecozzi^11^, Chamseddine Mediouni^10^, Bruno Megarbane^7^, Mohamed Meghadecha^22^, Élodie Mejean^12^, Arsene Mekinian^4^, Nour Mekki Abdelhadi^6^, Rania Mekni^3^, Thinhinan Sabrina Meliti^3^, Breno Melo Lima^18^, Paris Meng^12^, Soraya Merbah^3^, Fadhila Messani^2^, Yasmine Messaoudi^3^, Baboo-Irwinsingh Mewasing^12^, Lydia Meziane^3^, Carole Michelot-Burger^11^, Françoise Mignot^18^, Fadi Hillary Minka^7^, Makoto Miyara^3^, Pierre Moine^15^, Jean-Michel Molina^18^, Anaïs Montegnies-Boulet^5^, Alexandra Monti^21^, Claire Montlahuc^18^, Anne-Lise Montout^3^, Alexandre Moores^5^, Caroline Morbieu^5^, Helene Mortelette^14^, Stéphane Mouly^7^, Rosita Muzaffar^18^, Cherifa Iness Nacerddine^3^, Marine Nadal^12^, Hajer Nadif^3^, Kladoum Nassarmadji^7^, Pierre Natella^17^, Sandrine Ndingamondze^3^, Stefan Neraal^5^, Caroline Nguyen^6^, Bao N’Guyen^3^, Isabelle Nion Larmurier^4^, Luc Nlomenyengue^14^, Nicolas Noel^9^, Hilario Nunes^2^, Edris Omar^3^, Zineb Ouazene^4^, Elise Ouedraogo^2^, Wassila Ouelaa^3^, Anissa Oukhedouma^3^, Yasmina Ould Amara^3^, Herve Oya^3^, Johanna Oziel^2^, Thomas Padilla^3^, Elena Paillaud^26^, Solenne Paiva^3^, Beatrice Parfait^5^, Perrine Parize^11^, Christophe Parizot^3^, Antoine Parrot^12^, Arthur Pavot^9^, Laetitia Peaudecerf^5^, Frédéric Pene^5^, Marion Pepin^10^, Julie Pernet^3^, Claire Pernin^7^, Mylène Petit^2^, Olivier Peyrony^18^, Marie-Pierre Pietri^22^, Olivia Pietri^4^, Marc Pineton De Chambrun^3^, Michelle Pinson^13^, Claire Pintado^18^, Valentine Piquard^6^, Christine Pires^3^, Benjamin Planquette^14^, Sandrine Poirier^8^, Anne-Laure Pomel^8^, Stéphanie Pons^3^, Diane Ponscarme^18^, Annegaelle Pourcelot^9^, Valérie Pourcher^3^, Anne Pouvaret^11^, Florian Prever^4^, Miresta Previlon^18^, Margot Prevost^3^, Marie-Julie Provoost^7^, Cyril Quemeneur^3^, Cédric Rafat^12^, Agathe Rami^7^, Brigitte Ranque^14^, Maurice Raphael^9^, Jean Herle Raphalen^11^, Anna Rastoin^7^, Mathieu Raux^3^, Amani Rebai^2^, Michael Reby^25^, Alexis Regent^5^, Asma Regrag^14^, Matthieu Resche-Rigon^18^, Quentin Ressaire^18^, Christian Richard^9^, Mariecaroline Richard^3^, Maxence Robert^3^, Benjamin Rohaut^3^, Camille Rolland-Debord^12^, Jacques Ropers^3^, Anne-Marie Roque-Afonso^24^, Charlotte Rosso^30^, Mélanie Rousseaux^4^, Nabila Rousseaux^3^, Swasti Roux^26^, Lorène Roux^4^, Claire Rouzaud^11^, Antoine Rozes^3^, Emma Rubenstein^7^, Jean-Marc Sabate^2^, Sheila Sabet^12^, Sophie-Caroline Sacleux^24^, Nathalie Saidenberg Kermanach^2^, Faouzi Saliba^24^, Dominique Salmon^22^, Laurent Savale^31^, Guillaume Savary^3^, Rebecca Sberro^11^, Anne Scemla^11^, Frederic Schlemmer^17^, Mathieu Schwartz^7^, Saïd Sedfi^3^, Samia Sefir-Kribel^5^, Philippe Seksik^4^, Pierre Sellier^7^, Agathe Selves^3^, Nicole Sembach^14^, Luca Semerano^2^, Marie-Victoire Senat^9^, Damien Sene^7^, Alexandra Serris^11^, Lucile Sese^2^, Naima Sghiouar^15^, Johanna Sigaux^2^, Martin Siguier^12^, Johanne Silvain^3^, Noémie Simon^3^, Tabassome Simon^4^, Lina Innes Skandri^2^, Miassa Slimani^2^, Aurélie Snauwaert^6^, Harry Sokol^4^, Heithem Soliman^4^, Nisrine Soltani^9^, Benjamin Soyer^7^, Gabriel Steg^6^, Lydia Suarez^7^, Tali-Anne Szwebel^5^, Kossi Taffame^3^, Yacine Tandjaoui-Lambiotte^2^, Claire Tantet^2^, Mariagrazia Tateo^18^, Igor Theodose^18^, Pierre clement Thiebaud^4^, Caroline Thomas^4^, Kelly Tiercelet^18^, Julie Tisserand^9^, Carole Tomczak^18^, Krystel Torelino^3^, Fatima Touam-Ext^11^, Lilia Toumi^11^, Gustave Toury^14^, Mireille Toy-Miou^3^, Olivia Tran Dinh Thanh Lien^7^, Alexy Trandinh^6^, Jean-Marc Treluyer^5^, Baptiste Trinque^7^, Jennifer Truchot^5^, Florence Tubach^3^, Sarah Tubiana^6^, Simone Tunesi^19^, Matthieu Turpin^12^, Agathe Turpin^3^, Tomas Urbina^4^, Rafael Usubillaga Narvaez^22^, Yurdagul Uzunhan^2^, Prabakar Vaittinadaayar^27^, Arnaud Valent^18^, Maelle Valentian^12^, Nadia Valin^4^, Hélène Vallet^4^, Marina Vaz^3^, Miguel-Alejandro Vazquezibarra^7^, Benoit Vedie^14^, Laetitia Velly^3^, Celine Verstuyft^9^, Cedric Viallette^3^, Eric Vicaut^7^, Dorothee Vignes^8^, Damien Vimpere^11^, Myriam Virlouvet^9^, Guillaume Voiriot^12^, Lena Voisot^21^, Emmanuel Weiss^27^, Nicolas Weiss^3^, Anaïs Winchenne^2^, Youri Yordanov^4^, Lara Zafrani^18^, Mohamad Zaidan^9^, Wissem Zaidi^4^, Cathia Zak^12^, Aida Zarhrate-Ghoul^3^, Ouassila Zatout^6^, Suzanne Zeino^9^, Michel Zeitouni^3^, Naïma Zemirli^3^, Lorene Zerah^3^, Ounsa Zia^3^, Marianne Ziol^19^, Oceane Zolario^4^, Julien Zuber^11^

^1^DRCI-APHP, Paris, France, ^2^Hôpital Avicenne, Bobigny, France, ^3^Hôpital Pitié-Salpêtrière, Paris, France, ^4^Hôpital Saint-Antoine, Paris, France, ^5^Hôpital Cochin, Paris, France, ^6^Hôpital Bichat, Paris, France, ^7^Hôpital Lariboisière, Paris, France, ^8^Hôpital Antoine Béclère, Clamart, France, ^9^Hôpital Kremlin Bicêtre, Le Kremlin-Bicêtre, France, ^10^Hôpital Ambroise-Paré, Boulogne Billancourt, France, ^11^Hopital Necker Enfants malades, Paris, France, ^12^Hôpital Tenon, Paris, France, ^13^Hôpital Louis Mourier, Colombes, France, ^14^Hôpital Européen Georges Pompidou, Paris, France, ^15^Hôpital Raymond Poincaré, Garches, France, ^16^Hôpital Antoine Béclère, Calmart, France, ^17^Hôpital Henri Mondor, Créteil, France, ^18^Hôpital Saint Louis, Paris, France, ^19^Hôpital Jean Verdier, Bondy, France, ^20^Université Paris-Sorbonne, Hôpital Pitié-Salpêtrière, INSERM, Paris, France, ^21^Hôpital Charles Foix, Ivry-sur-Seine, France, ^22^Hôpital Hôtel Dieu, Paris, France, ^23^Hôpital Broca, Paris, France, ^24^Hôpital Paul-Brousse, Villejuif, France, ^25^Hôpital Rothschild, Paris, France, ^26^Hôpital Corentin Celton, Issy-les-Moulineaux, France, ^27^Hôpital Beaujon, Clichy, France, ^28^Hôpital Albert Chenevier, Créteil, France, ^29^Hôpital Sainte-Périne, Paris, France, ^30^Université Paris-Sorbonne, Hôpital Pitié-Salpêtrière, INSERM, CNRS, Paris, France, ^31^Université Paris-Saclay, Hôpital Kremlin Bicêtre, INSERM, Le Kremlin-Bicêtre, France

***Members of Amsterdam UMC Covid-19 Biobank*:** Michiel van Agtmael^2^, Anne Geke Algera^1^, Brent Appelman^2^, Frank van Baarle^1^, Diane Bax^3^, Martijn Beudel^4^, Harm Jan Bogaard^5^, Marije Bomers^2^, Peter Bonta^5^, Lieuwe Bos^1^, Michela Botta^1^, Justin de Brabander^2^, Godelieve de Bree^2^, Sanne de Bruin^1^, David T. P. Buis^1^, Marianna Bugiani^5^, Esther Bulle^1^, Osoul Chouchane^2^, Alex Cloherty^3^, Mirjam Dijkstra^12^, Dave A. Dongelmans^1^, Romein W. G. Dujardin^1^, Paul Elbers^1^, Lucas Fleuren^1^, Suzanne Geerlings^2^, Theo Geijtenbeek^3^, Armand Girbes^1^, Bram Goorhuis^2^, Martin P. Grobusch^2^, Florianne Hafkamp^3^, Laura Hagens^1^, Jorg Hamann^7^, Vanessa Harris^2^, Robert Hemke^8^, Sabine M. Hermans^2^, Leo Heunks^1^, Markus Hollmann^6^, Janneke Horn^1^, Joppe W. Hovius^2^, Menno D. de Jong^9^, Rutger Koning^4^, Endry H. T. Lim^1^, Niels van Mourik^1^, Jeaninne Nellen^2^, Esther J. Nossent^5^, Frederique Paulus^1^, Edgar Peters^2^, Dan A. I. Pina-Fuentes^4^, Tom van der Poll^2^, Bennedikt Preckel^6^, Jan M. Prins^2^, Jorinde Raasveld^1^, Tom Reijnders^2^, Maurits C. F. J. de Rotte^12^, Michiel Schinkel^2^, Marcus J. Schultz^1^, Femke A. P. Schrauwen^12^, Alex Schuurmans^10^, Jaap Schuurmans^1^, Kim Sigaloff^1^, Marleen A. Slim^1,2^, Patrick Smeele^5^, Marry Smit^1^, Cornelis S. Stijnis^2^, Willemke Stilma^1^, Charlotte Teunissen^11^, Patrick Thoral^1^, Anissa M. Tsonas^1^, Pieter R. Tuinman^2^, Marc van der Valk^2^, Denise P. Veelo^6^, Carolien Volleman^1^, Heder de Vries^1^, Lonneke A. Vught^1,2^, Michèle van Vugt^2^, Dorien Wouters^12^, A. H. (Koos) Zwinderman^13^, Matthijs C. Brouwer^4^, W. Joost Wiersinga^2^, Alexander P. J. Vlaar^1^, Diederik van de Beek^4^

^1^Department of Intensive Care, Amsterdam UMC, Amsterdam, Netherlands. ^2^Department of Infectious Diseases, Amsterdam UMC, Amsterdam, Netherlands. ^3^Experimental Immunology, Amsterdam UMC, Amsterdam, Netherlands. ^4^Department of Neurology, Amsterdam UMC, Amsterdam Neuroscience, Amsterdam, Netherlands. ^5^Department of Pulmonology, Amsterdam UMC, Amsterdam, Netherlands. ^6^Department of Anesthesiology, Amsterdam UMC, Amsterdam, Netherlands. ^7^Amsterdam UMC Biobank Core Facility, Amsterdam UMC, Amsterdam, Netherlands. ^8^Department of Radiology, Amsterdam UMC, Amsterdam, Netherlands. ^9^Department of Medical Microbiology, Amsterdam UMC, Amsterdam, Netherlands. ^10^Department of Internal Medicine, Amsterdam UMC, Amsterdam, Netherlands. ^11^Neurochemical Laboratory, Amsterdam UMC, Amsterdam, Netherlands. ^12^Department of Clinical Chemistry, Amsterdam UMC, Amsterdam, Netherlands. ^13^Department of Clinical Epidemiology, Biostatistics and Bioinformatics, Amsterdam UMC, Amsterdam, Netherlands.

***Members of NIAID-USUHS COVID Study Group*:** Miranda F. Tompkins^1^, Camille Alba^1^, Andrew L. Snow^2^, Daniel N. Hupalo^1^, John Rosenberger^1^, Gauthaman Sukumar^1^, Matthew D. Wilkerson^1^, Xijun Zhang^1^, Justin Lack^3^, Andrew J. Oler^4^, Kerry Dobbs^5^, Ottavia M. Delmonte^5^, Jeffrey J. Danielson^5^, Andrea Biondi^6^, Laura Rachele Bettini^6^, Mariella D’Angiò^6^, Ilaria Beretta^7^, Luisa Imberti^8^, Alessandra Sottini^8^, Virginia Quaresima^8^, Eugenia Quiros-Roldan^9^, Camillo Rossi^10^, Riccardo Castagnoli^11^, Daniela Montagna^12^, Luigi D. Notarangelo^13^.

^1^American Genome Center, Uniformed Services University of the Health Sciences, Bethesda, MD, USA; Henry M. Jackson Foundation for the Advancement of Military Medicine, Bethesda, MD, USA. ^2^Department of Pharmacology and Molecular Therapeutics, Uniformed Services University of the Health Sciences, Bethesda, MD, USA. ^3^NIAID Collaborative Bioinformatics Resource, Frederick National Laboratory for Cancer Research, Leidos Biomedical Research Inc., Frederick, MD, USA. ^4^Bioinformatics and Computational Biosciences Branch, Office of Cyber Infrastructure and Computational Biology, NIAID, NIH, Bethesda, MD, USA. ^5^Laboratory of Clinical Immunology and Microbiology, Division of Intramural Research, NIAID, NIH, Bethesda, MD, USA. ^6^Pediatric Departement and Centro Tettamanti-European Reference Network PaedCan, EuroBloodNet, MetabERN-University of Milano-Bicocca-Fondazione MBBM-Ospedale, San Gerardo, Monza, Italy. ^7^Department of Infectious Diseases, University of Milano-Bicocca, San Gerardo Hospital, Monza, Italy. ^8^CREA Laboratory, Diagnostic Department, ASST Spedali Civili di Brescia, Brescia, Italy. ^9^Department of Infectious and Tropical Diseases, University of Brescia and ASST Spedali Civili di Brescia, Brescia, Italy. ^10^Chief Medical Officer, ASST Spedali Civili di Brescia, Brescia, Italy. ^11^Department of Pediatrics, Fondazione IRCCS Policlinico San Matteo, University of Pavia, Pavia, Italy. ^12^Laboratory of Immunology and Transplantation, Fondazione IRCCS Policlinico San Matteo, Pavia, Italy; Department of Clinical, Surgical, Diagnostic and Pediatric Sciences, University of Pavia, Pavia, Italy. ^13^National Institute of Allergy and Infectious Diseases, National Institutes of Health, Bethesda, MD, USA.

## Web resources

HLA alleles distribution across populations:

Allele frequency Net Database: https://github.com/slowkow/allelefrequencies

1000 Genomes Project: http://ftp.1000genomes.ebi.ac.uk/vol1/ftp/data_collections/HLA_types/

## Data availability

Data supporting the findings of this study are available within the manuscript and supplemental files. The whole-genome sequencing data of anonymized patients recruited through the National Institutes of Health (NIH) and sequenced at the National Institute of Allergy and Infectious Diseases (NIAID) through the Uniformed Services University of the Health Sciences (USUHS)/the American Genome Center (TAGC) are available under dbGaP submission phs002245.v1. Other patients were not consented to share the raw WES/WGS data files beyond the research and clinical teams.

## Author contributions

AM, A Cobat, and AB performed computational analysis. AM, ETC, IN, EB, RT, KMSB, YZ, INB, MK, A Catchpole, JBL, CLD, VSS, A Cobat and AB performed or supervised experiments, generated and analyzed data, and contributed to the manuscript by providing figures and tables. SGT, ANS, JG, CB, GG, FT, PH, SYZ, QZ, CC, JF, JJG, VSS and the consortium collaborators evaluated and recruited patients and /or controls. AM, LA, JLC, A Cobat, and AB wrote the manuscript. CC, JJG, LA, JLC, A Cobat, and AB supervised the project. All authors edited the manuscript. All authors read and approved the final manuscript.

## Notes

### Funding Statement

Funding was provided to the Desert Research Institute (DRI) by the Nevada Governors Office of Economic Development. Funding was provided by Renown Health and the Renown Health Foundation.
The Laboratory of Human Genetics of Infectious Diseases is supported by the Howard Hughes Medical Institute, the Rockefeller University, the St. Giles Foundation, the National Institutes of Health (NIH) (R01AI63029), the National Center for Advancing Translational Sciences (NCATS), NIH Clinical and Translational Science Award (CTSA) program (UL1 TR001866), the Yale Center for Mendelian Genomics and the GSP Coordinating Center funded by the National Human Genome Research Institute (NHGRI) (UM1HG006504 and U24HG008956), the Yale High Performance Computing Center (S10OD018521), the Fisher Center for Alzheimers Research Foundation, the JPB Foundation, the Meyer Foundation, the French National Research Agency (ANR) under the Investments for the Future program (ANR-10-IAHU-01), the Integrative Biology of Emerging Infectious Diseases Laboratory of Excellence (ANR-10-LABX-62-IBEID), the French Foundation for Medical Research (FRM) (EQU201903007798), the ANRS-COV05, ANR GENVIR (ANR-20-CE93-003), and ANR AI2D (ANR-22-CE15-0046) projects, the ANR-RHU program (ANR-21-RHUS-08), the European Union Horizon 2020 research and innovation program under grant agreement No. 824110 (EASI-genomics), the HORIZON-HLTH-2021-DISEASE-04 program under grant agreement 01057100 (UNDINE), the Square Foundation, Grandir - Fonds de solidarite pour lenfance, Fondation du Souffle, the SCOR Corporate Foundation for Science, William E. Ford, General Atlantic Chairman and Chief Executive Officer, Gabriel Caillaux, General Atlantic Co-President, Managing Director and Head of business in EMEA, and the General Atlantic Foundation, the Battersea & Bowery Advisory Group, The French Ministry of Higher Education, Research, and Innovation (MESRI-COVID-19), Institut National de la Sante et de la Recherche Medicale (INSERM), REACTing-INSERM and the University of Paris Cite. A.N.S. is supported by European Union Horizon Health research and innovation program under grant agreement No 101057100, project UNDINE.
The study was supported by the ORCHESTRA project, which has received funding from the European Union Horizon 2020 research and innovation program under grant agreement No 10101616. The French COVID Cohort study group was sponsored by INSERM and supported by the REACTing consortium and by a grant from the French Ministry of Health (Grant PHRC 20-0424). The Cov-Contact Cohort was supported by the REACTing consortium, the French Ministry of Health, and the European Commission (Grant RECOVER WP 6). The COVIDeF study group was supported by the French Ministry of Health, Fondation AP-HP et Programme Hospitalier de Recherche Clinique (PHRC COVID-19-20-0048). Y.Z. and H.C.S. are supported by the Intramural Research Program of the National Institute of Allergy and Infectious Diseases, NIH. G.N. and A.N. are supported by Regione Lazio (Research Group Projects 2020) No. A0375-2020-36663, GecoBiomark. This project has received funding from the European Research Council (ERC) under the European Union Horizon 2020 research and innovation program (grant agreement no. 948959). This work is supported by the Swiss National Science Foundation (grant # 310030L_197721 to JF). This work is supported by ERN-RITA. The Canarian Sequencing Hub is funded by Instituto de Salud Carlos III (COV20_01333, and COV20_01334, and PI20/00876) and Spanish Ministry of Science and Innovation (RTC-2017-6471-1; AEI/FEDER, UE), co-financed by the European Regional Development Funds, A way of making Europe, from the European Union, and Cabildo Insular de Tenerife (CGIEU0000219140 and Apuestas cientificas del ITER para colaborar en la lucha contra la COVID-19). This work was funded, at least in part, by grants AJF202019 and AJF20259 from Al Jalila Foundation, Dubai, United Arab Emirates. Sample processing at IrsiCaixa was possible thanks to the crowdfunding initiative YoMeCorono.

### Author Declarations

All the enrolled participants provided written informed consent for participation and were recruited through protocols conforming to local ethics requirements. For patients enrolled in the Helix DNA Discovery Project, ethics approval was reviewed and obtained from the Western Institutional Review Board. For patients enrolled in the Healthy Nevada Project (HNP), the University of Nevada, Reno Institutional Review Board approved the study (project 956068-12). For patients enrolled in the French COVID cohort (ClinicalTrials.gov NCT04262921), ethics approval was obtained from the Comite de Protection des Personnes Ile De France VI (ID RCB, 2020-A00256-33) or the Ethics Committee of Erasme Hospital (P2020/203). For participants enrolled in the COV-Contact study (ClinicalTrials.gov NCT04259892), ethics approval was obtained from the CPP IDF VI (ID RCB, 2020-A00280-39). For patients enrolled in the Italian cohort, ethics approval was obtained from the University of Milano-Bicocca School of Medicine, San Gerardo Hospital, Monza-Ethics Committee of the National Institute of Infectious Diseases Lazzaro Spallanzani (84/2020) (Italy), and the Comitato Etico Provinciale (NP 4000-Studio CORONAlab). STORM-Health care workers were enrolled in the STudio OsseRvazionale sullo screening dei lavoratori ospedalieri per COVID-19 (STORM-HCW) study, with approval from the local institutional review board (IRB) obtained on June 18, 2020. Patients and relatives from San Raffaele Hospital (Milan) were enrolled in COVID-BioB/Gene-COVID protocols and, for additional studies, TIGET-06, with the approval of the local ethics committee. Patients and relatives from Rome were enrolled in Protocol no. 50/20 (Tor Vergata University Hospital). Informed consent was obtained from each patient. For the patients enrolled in the COVIDeF Study Group (ClinicalTrials.gov NCT04352348), ethics approval was obtained from the Comite de Protection des Personnes Ile de France XI (ID RCB, 2020-A00754-35). For patients enrolled in Spain, the study was approved by the Committee for Ethical Research of the Infanta Leonor University Hospital, code 008-20; the Committee for Ethical Research of the 12 de Octubre University Hospital, code 16/368; the Bellvitge University Hospital, code PR127/20; the University Hospital of Gran Canaria Dr. Negrin, code 2020-200-1 COVID-19; and the Vall d Hebron University Hospital, code PR(AMI)388/2016. Anonymized samples were sequenced at the National Institute of Allergy and Infectious Diseases (NIAID) through the Uniformed Services University of the Health Sciences(USUHS)/the American Genome Center (TAGC) under nonhuman subject research conditions; no additional IRB consent was required at the National Institutes of Health (NIH). For patients enrolled in the Swedish COVID cohort, ethics approval was obtained from the Swedish Ethical Review Agency (2020-01911 05). For patients enrolled in the SARS-CoV-2 Human Challenge Characterisation Study, ethics approval was obtained from the UK Health Research Authority Ad Hoc Specialist Ethics Committee (reference: 20/UK/0002). Written informed consent was obtained from participants before screening and enrollment.

