## Supplementary Figures for "Lack of association between HLA and asymptomatic SARS-CoV-2 infection"

### Figure S1

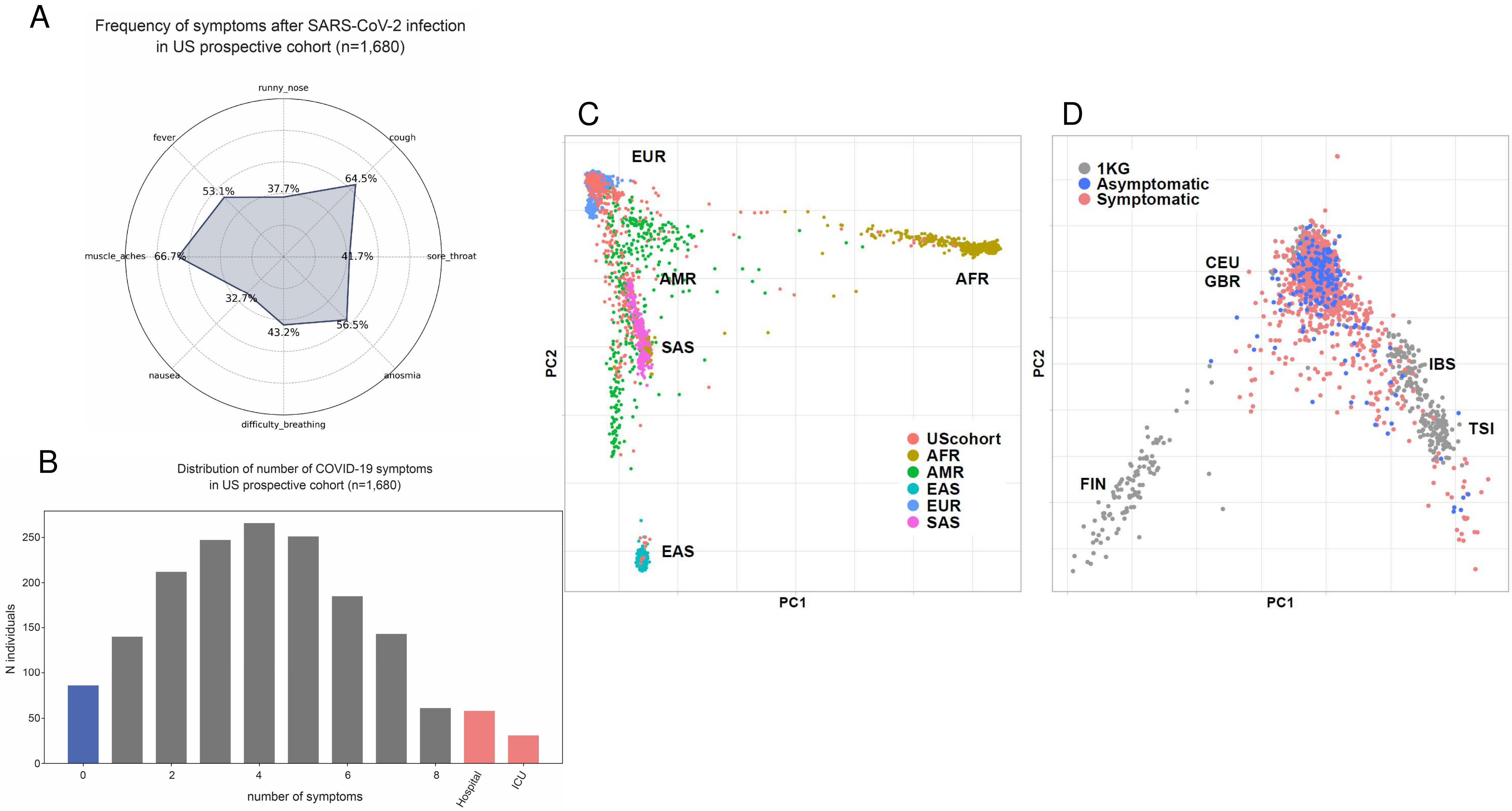


**Figure S1: COVID-19 symptoms in a US prospective cohort.**

A: Radar plot showing the % of infected individuals reporting each symptom.

B: Distribution of the number of symptoms per individual infected with SARS-CoV-2. Patients who had to be hospitalized or admitted to the ICU are represented separately (pink bars). Infected individuals who reporting no symptoms are represented by the blue bar.

C: PCA plot displaying the US prospective cohort (orange dots) overlapped with 1000 Genomes Project samples labeled with their known ancestry (AFR: African, AMR: American, EAS: East Asian, EUR: European, SAS: South Asian).

D: PCA plot displaying the European genetic ancestry subsets of the US prospective cohort and from 1000 Genomes Projects. Origin of 1000 Genomes Project (1KG) samples are indicated (CEU: Utah residents with Northern and Western European ancestry, FIN: Finnish in Finland, GBR: British in England and Scotland, IBS: Iberian population in Spain, TSI: Toscani in Italy).

### Figure S2

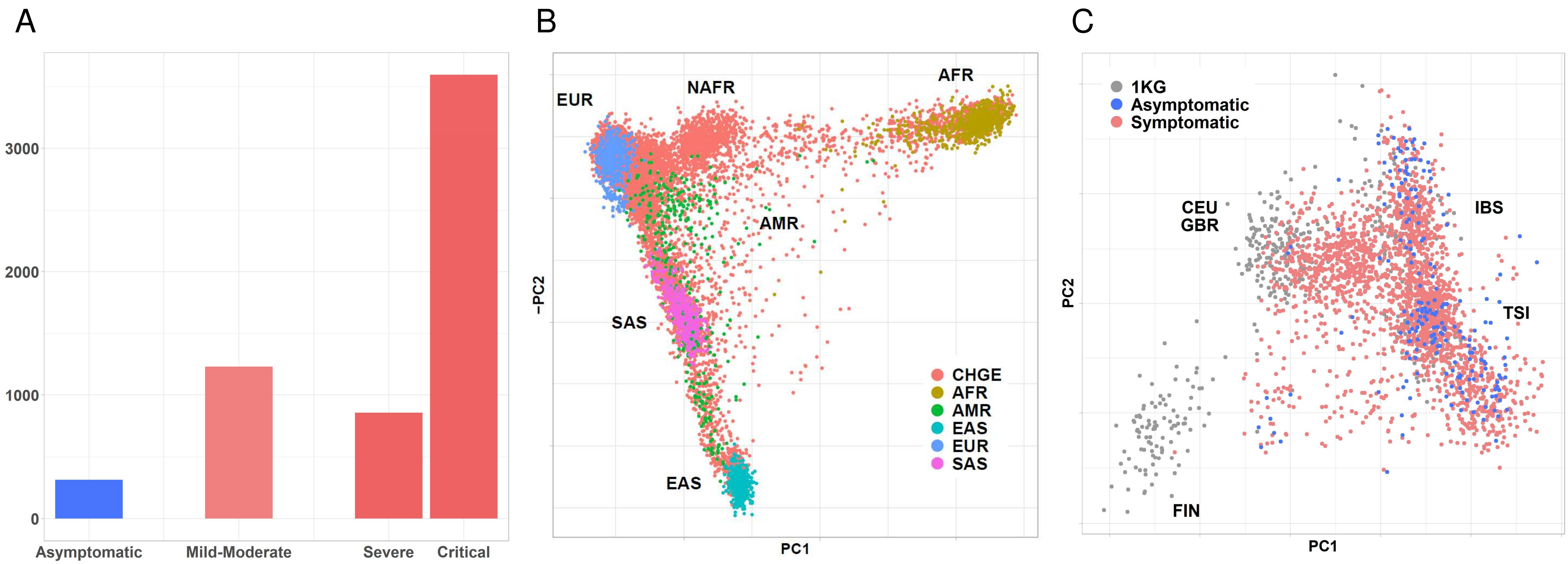


**Figure S2: COVID-19 symptoms in the CHGE cohort.**
A: Distribution of the number of individuals per category of severity.
B: PCA plot displaying the CHGE cohort (orange dots) overlapped with the samples from 1000 Genomes Project. Ancestry of 100 Genomes Project samples are indicated (AFR: African, AMR: American, EAS: East Asian, EUR: European, NAFR: North African, SAS: South Asian).
C: PCA plot displaying the European genetic ancestry subsets of the CHGE cohort and from 1000 Genomes Projects. Origin of 1000 Genomes Project (1KG) samples are indicated (CEU: Utah residents with Northern and Western European ancestry, FIN: Finnish in Finland, GBR: British in England and Scotland, IBS: Iberian population in Spain, TSI: Toscani in Italy).

Figure S3

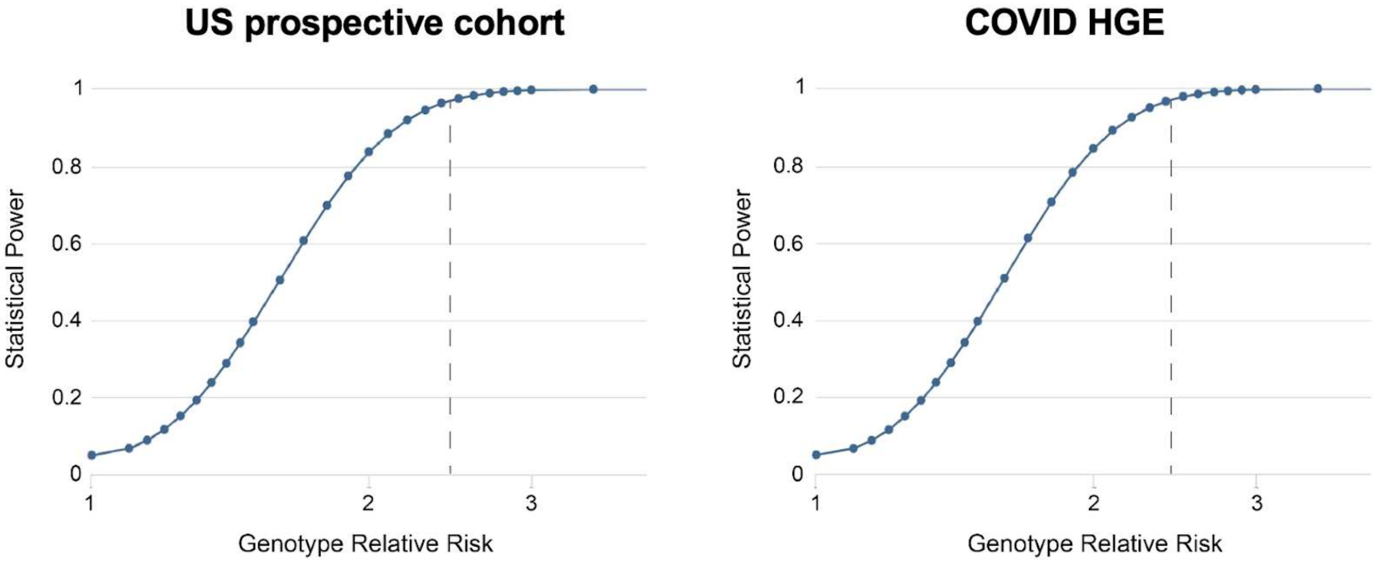


**Figure S3: Power curves.**
Statistical power as a function of genotype relative risk, calculated under a dominant inheritance model, with a HLA-B*15:01 frequency of 0.05, a prevalence of asymptomatic infection of 0.1, and a p-value threshold of 0.05. Curves plotted with the Genetic Association Study Power Calculator (https://csg.sph.umich.edu/abecasis/gas_power_calculator). The dotted line represent the Odds ratio obtained by Augusto, Murdolo & Chatzileontiadou et al. (OR=2.4).

Figure S4

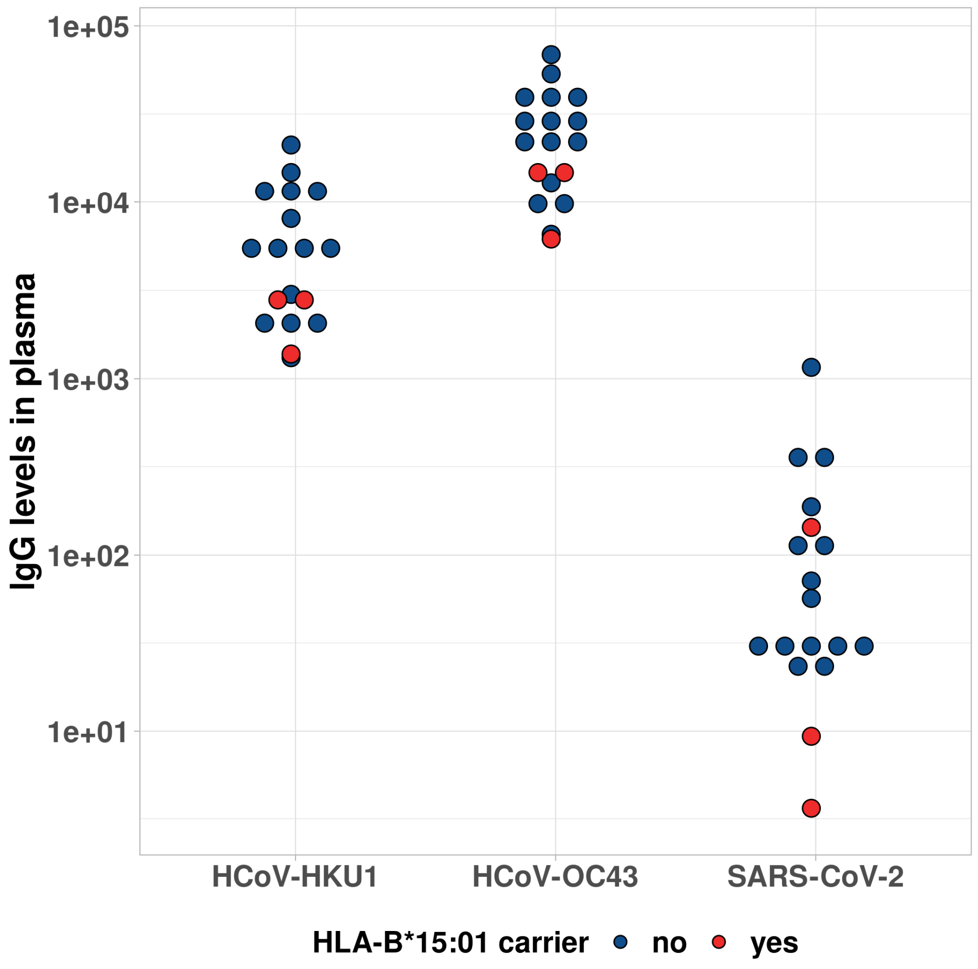


**Figure S4: Seasonal CoV antibodies in the SARS-CoV-2 Human Challenge Characterisation Study.**

Plasma IgG was quantified in baseline samples from the SARS-CoV-2 human challenge characterisation study participants (Infected, n=17) for HKU1-CoV Spike protein, OC43-CoV Spike protein and SARS-CoV-2 Spike protein as a negative control. Carriers of HLA-B*15:01 are indicated in red. Arbitrary units per milliliter.

Figure S5

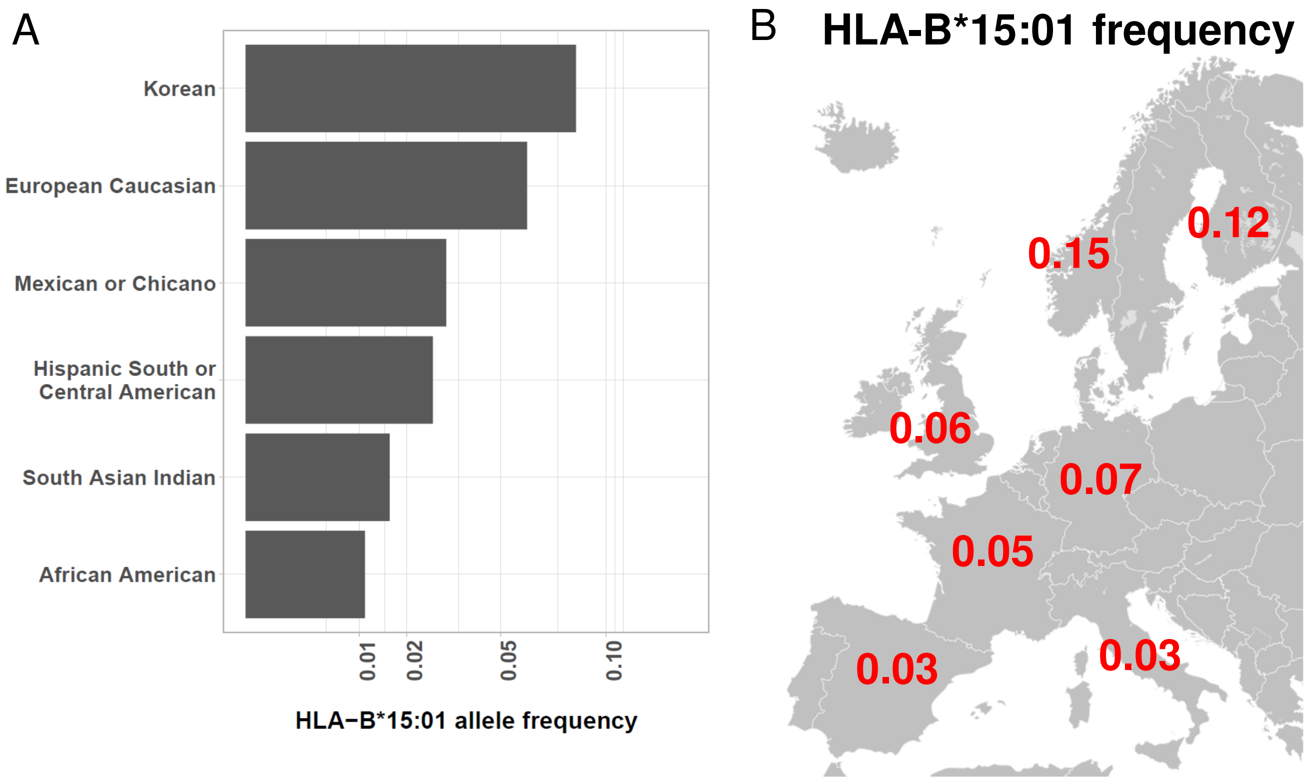


**Figure S5: Frequency of HLA-B*15:01 allele** (A): for the various subpopulations in the US and (B) in Europe. Data from Allele Frequency Net Database (www.allelefrequencies.net) and 1000 Genomes Project.
